# Genetic and environmental risk factors for intracranial aneurysm and subarachnoid haemorrhage among patients with Autosomal Dominant Polycystic Kidney Disease

**DOI:** 10.64898/2026.08.07.26359892

**Authors:** Lea Urpa, Shohdan Osman, Tomas Visser, Omid Sadeghi Alavijeh, Ashwini Shanmugam, Winston Fung, Elhussein Elhassan, Omri Teltsh, Aleksanteri Asikainen, Edmund Gilbert, Gianpiero L. Cavalleri, Kuruvilla Sebastian, Shane Lavin, Rodosthenis S. Rodosthenous, Karol Estrada, Rahul Raj, Aarno Palotie, Teemu Niiranen, Umberto Simola, Juha Martola, Mohsen Javadpour, Patrick Nicholson, Mikka Korja, Daniel P. Gale, Patrik Finne, Peter J. Conlon, Daniel Gordin

## Abstract

Intracranial aneurysms (IA) and their rupture (subarachnoid haemorrhage, SAH) are a rare but serious complication of autosomal dominant polycystic kidney disease (ADPKD). Both genetic and environmental risk factors contribute to the pathogenesis of IA and SAH, but specific information on risk factors in the ADPKD population are limited and international clinical guidelines mainly recommend screening for ADPKD patients with a family history of IA or SAH. We assessed the associations of monogenic variants, polygenic risk, and clinical factors with the diagnosis of IA or SAH in 2,200 adult ADPKD patients from three cohorts: the Irish Kidney Gene Project (IKGP, n=475), FinnGen (n=826), and Genomics England (GEL, n=899). Polygenic risk scores (PRS) for IA, hypertension, and smoking intensity were derived from previously published GWAS summary statistics and additionally conditioned using mtCOJO to account for their genetic correlation. IA or SAH was diagnosed in 158 of 2,200 patients (7.2%; mean age at diagnosis 51.1 years). Female sex (HR 2.21, 95% CI 1.50-3.25, p=6.52×10^−5^) and smoking (HR 2.06, 95% CI 1.43-2.96, p=9.49 x10^−5^) were associated with IA or SAH in univariate Cox proportional hazards models, while diabetes was protective (HR 0.39, 95% CI 0.22-0.70, p=1.37×10^−3^). Monogenic variant status was not found to associate with increased risk of IA or SAH. Polygenic score for IA was associated with increased risk of IA or SAH, with a 1 standard deviation increase in PRS associated with a pooled hazard ratio (HR) of 1.30 (95% CI 1.09–1.57, p=4.66×10^−3^), and the association of the conditioned IA PRS remained in multivariate Cox proportional hazards models accounting for clinical factors and monogenic variants (HR 1.26, 95% CI 1.03–1.55, p=0.02). The addition of polygenic scores added significant prognostic information for IA or SAH beyond clinical risk factors in FinnGen (p=3.61×10⁻^3^) and GEL (p=8.84×10⁻^3^) but not in IKGP, as assessed by the likelihood ratio test. Overall, our study shows that the strongest risk factors for IA or SAH in ADPKD patients are smoking history, female sex, and polygenic risk for IA. Hypertension was not associated with IA (HR 0.50, 95% CI 0.24-1.00, p=0.051), likely due to the high prevalence of hypertension in this population across cohorts (69.6-85.3%). While family history may be considered a proxy of genetic risk, this study suggests that family history itself may be of limited utility in discriminating individuals with ADPKD at risk of IA or SAH.

## Introduction

Autosomal Dominant Polycystic Kidney Disease (ADPKD) is one of the most serious inherited renal disorders and a major cause of kidney failure (KF)^1,2^. Intracranial aneurysms (IA) are significant extrarenal manifestations of ADPKD, with prevalence estimates ranging from 8% to 20% in ADPKD patients compared to 2-3% in the general population^2–5^ but even up to 10% in postmenopausal female smokers^6^. Aneurysmal subarachnoid haemorrhage (SAH) occurs at a higher rate in ADPKD than in the general population (0.57 vs 0.079 per 1000 person-years)^7–9^, with case fatality of approximately 30–40% and long-term cognitive or functional impairment in many survivors^10,11^, although absolute risk remains small.

Current clinical practice recommends targeting screening mainly to individuals with a personal or family history of IA or SAH^5,12–14^, yet most SAHs occur in patients without family history^3^. Family history can be considered a proxy and inexpensive alternative to genetic risk profiling, yet it captures both shared environmental and inherited genetic risk factors. In non-ADPKD patients, the estimated heritability of SAH is 41%, based on a large multinational twin study^15^. This inherited genetic risk is also composed of both common genetic variants that cumulatively impact disease risk, which can be assessed by polygenic risk scores^16–21^, and rare genetic variation unique to the family. In fact, polygenic risk has been shown to be complementary to family history^22^, and individuals within a family may have differing levels of risk depending on the variants they inherit^23^, suggesting that family history may be of limited utility as a prognostic factor for individual patients.

Concerning rare genetic variation, the contribution of germline ADPKD variants to risk of IA or SAH is still incompletely understood, both from the perspective of the gene involved (*PKD1, PKD2,* or other rarer causative genes) and the variant class (truncating versus non-truncating). Previous work has shown that IA was diagnosed more than twice as often in carriers of *PKD1* variants compared with *PKD2* carriers, although neither variant type nor position within *PKD1* showed a clear effect^24^, and a smaller single-centre study reported that IAs developed earlier in carriers of *PKD1* truncating variants than in those with non-truncating substitutions^25^. Beyond ADPKD, population-level analyses indicate that rare damaging *PKD1* variants also raise IA risk in the general population^26^.

Common genetic variation has also been associated with IA risk in the general population^27,28^, and recent work by Bakker et al^27^ developed an IA-specific PRS that demonstrated robust risk stratification in general population cohorts, with individuals in the highest PRS decile showing significantly elevated IA risk compared to those with average genetic burden. This genetic risk for IA overlapped with genetic predisposition to smoking intensity and high blood pressure, two of the most important modifiable IA risk factors in the general population, with Mendelian randomisation further supporting a causal role for both on IA risk^27^. However, it is unclear whether polygenic risk scores capturing common-variant IA risk are associated with IA or SAH in an ADPKD population, and how this risk interacts with the underlying monogenic ADPKD variants and other clinical risk factors for IA or SAH.

Overall, longitudinal and well-characterized studies including both genetic and clinical data are lacking, and such studies may provide tools to improve optimal screening strategy for IA in ADPKD. Notably, screening practices vary considerably between countries and centres. The Kidney Disease: Improving Global Outcomes (KDIGO) 2025 ADPKD guideline acknowledges this uncertainty and identifies the frequency, risk stratification, and rescreening of IAs in ADPKD as priorities for future research^2^. With the aim of identifying patients with ADPKD who carry an elevated risk of not only IA but also SAH, we examined the joint contribution of polygenic risk, monogenic variants, and conventional clinical risk factors across three independent ADPKD populations.

## Results

### Study Population Characteristics

A total of 2,200 ADPKD patients were included in the analysis across three cohorts: 475 from the Irish Kidney Gene Project (IKGP), 826 from FinnGen, and 899 from Genomics England (GEL). Males comprised 47.5% of the total cohort, among individuals who developed IA or SAH, the mean age at diagnosis was 50.2 years in IKGP 51.6, years in FinnGen and 51.4 years in GEL, giving a pooled mean of 51.1 years (Table 1). Prevalence of SAH was comparable between cohorts, with 2.3% of IKGP ADPKD patients experiencing an episode of SAH, compared to 1.9% in FinnGen and 1.1% in GEL, but prevalence of IA differed markedly between the cohorts: 9.7% of IKGP patients experienced IA compared to 6.2% in FinnGen, with lower prevalence in Genomics England (2.7%). This difference in prevalence between cohorts is likely due to differences in diagnosis and screening across cohorts and scope of the source data. While SAH was well captured from direct access to clinical notes in IKGP and from pseudonymized electronic health records (EHR) in FinnGen, with more limited capture from pseudonymized EHR in GEL (see Methods), SAH is a significant and highly morbid clinical event. IA, however, requires screening for diagnosis (when not found incidentally via imaging), for which practices were markedly different across the cohorts: 53% of IKGP patients underwent screening for IA, compared to 16% in FinnGen. Due to the small absolute numbers of SAH across the three cohorts, we combined IA and SAH into a single endpoint for further epidemiological analyses.

**Table 1.** Characteristics of Study Participants across Three Cohorts.

| Characteristic | IKGP | FinnGen | GEL |
| --- | --- | --- | --- |
| Total sample size | 475 | 826 | 899 |
| Sex male | 222 (46.7%) | 397 (48.1%) | 425 (47.3%) |
| IA or SAH | 57 (12.0%) | 67 (8.1%) | 34 (3.8%) |
| IA | 46 (9.7%) | 51 (6.2%) | 24 (2.7%) |
| SAH | 11 (2.3%) | 16 (1.9%) | 10 (1.1%) |
| Mean age at IA or SAH diagnosis | 50.2 | 51.6 | 51.4 |
| Screened for aneurysm | 255 (53.6%) | 134 (16.2%) | 274(30.5%) |
| History of smoking | 51 (10.7%) | 406 (49.2%) | 101 (11.2%) |
| Imputed Smoking history | 92 (19.4%) | 235 (28.5%) | NA |
| Family history of IA or SAH | 128 (26.9%) | 120 (14.5%) | 13 (1.5%) |
| Dyslipidaemia | 172 (36.2%) | 163 (19.7%) | 113 (12.6%) |
| Kidney failure (KF) | 195 (41.1%) | 527 (63.8%) | 212 (23.6%) |
| Mean KF Age | 50.5 | 56.4 | 52.4 |
| Hypertension | 401 (84.4%) | 705 (85.3%) | 626 (69.6%) |
| Diabetes (T1D and T2D) | 40 (8.4%) | 249 (30.1%) | 91 (10.1%) |
| Rare variants assessed (n) | 475 (100%) | 617 (74.7%) | 899 (100%) |
| *PKD1* variant carrier | 334 (70.3%) | 325 (52.7%) | 514 (57.1%) |
| Truncating | 220 (46.3%) | 143 (23.2%) | 325 (36.1%) |
| Non-truncating | 105 (22.1%) | 182 (29.5%) | 189 (21.0%) |
| *PKD2* variant carrier | 36 (7.6%) | 39 (6.3%) | 120 (13.3%) |
| Truncating | 28 (5.9%) | 18 (2.9%) | 102 (11.3%) |
| Non-truncating | 8 (1.7%) | 21 (3.4%) | 18 (2.0%) |
| Other gene variant carrier | 11 (2.3%) | 22 (3.6%) | 53 (5.9%) |
| No variant found | 94 (19.8%) | 231 (37.4%) | 212 (23.6%) |

**Table1** presents the demographic and clinical characteristics of participants from three independent cohorts. The table demonstrates the distribution of key variables including intracranial aneurysm occurrence (IA), kidney failure status, and cardiovascular risk factors across the study populations. Notable differences exist in data completeness, particularly for smoking history and family history variables. IA = Intracranial aneurysm, SAH = subarachnoid haemorrhage, KF = Kidney Failure. Percentages for monogenic variant categories in FinnGen are calculated using the number of individuals for whom rare variants were assessed (n=617). Percentages for *PKD1* truncating vs non-truncating status are reported for individuals for which detailed *PKD1* information was known (n=325).

History of smoking also varied markedly between cohorts, with the highest rates of smoking in FinnGen (49.2%) compared to IKGP (10.7%) and in GEL (11.2%). This figure was likely influenced by the different ascertainment of smoking information in each cohort: in FinnGen smoking data was obtained through a combination of legacy survey data and information provided from biobanks, and the data providers note that smoking is enriched in the data as a single positive answer of having smoked at any point gives an ‘ever smoker’ value. In IKGP, smoking status was ascertained via clinical interview recorded in patient notes, while in GEL smoking status was ascertained via ICD codes in EHR (which is likely an underestimate of smoking status at recruitment).

Family history of IA or SAH also varied markedly across the cohorts, likely differing by definition of family history (see Methods for details). Rates ranged from 1.5% in GEL where reporting is based on ICD codes of family history of IA or SAH (also likely an underestimate of true family history status) to 26.9% in IKGP Of these, 43.0% had at least one affected first-degree relative, reporting is based on clinical interview as reported in medical records and the Progeny pedigree database. The ascertainment of family history of IA or SAH in FinnGen (14.5%) is notably different than the other two cohorts, as we utilized genetically inferred relatedness (reliable up to 3^rd^ degree relatives) and the same electronic health record-based ascertainment that was applied to cases, giving a strong reliability for family history.

Rates of dyslipidaemia (hypercholesterolemia, defined via ICD codes in FinnGen and GEL and via clinical notes in IKGP) were 12.6-36.2% across cohorts, with the highest being the Irish Kidney Gene Project. End-stage renal failure (kidney replacement therapy or dialysis) also varied markedly among cohorts, with lowest prevalence in GEL (23.6%) and highest in FinnGen (63.8%). Hypertension was high across all three cohorts, affecting 84.4% of individuals in IKGP, 85.3% in FinnGen, and 69.6% in GEL. Diabetes (Type 1 and Type 2) varied markedly among cohorts, 8.4% of ADPKD individuals affected in IKGP, 30.1% in FinnGen, and 10.1% in GEL. Monogenic variants in either *PKD1* or *PKD2* were identified in 77.9% of individuals in IKGP, 58.7% in FinnGen, and 70.4% in GEL. This may reflect differences in definition of likely pathogenic variants among cohorts, with research-based methods applied for FinnGen and GEL and clinical guidelines for IKGP (see Methods) but may also suggest undiscovered rare variants associated with polycystic kidney disease in the Finnish population.

### Intracranial aneurysm polygenic risk stratifies IA risk in ADPKD patients

Kaplan-Meier survival analysis revealed significant risk stratification across polygenic risk strata for IA PRS in a meta-analysis across cohorts (Figure 1). To disentangle the genetic predispositions to IA^27^, smoking^29^, and hypertension^30^, we utilized multi-trait conditional and joint analysis (mtCOJO)^31^ to condition the effect of genetic variants on IA after accounting for their effect on hypertension and smoking intensity (see Methods). These conditioned GWAS summary statistics were then used to generate polygenic risk scores (Figure 1, top row) and compared to polygenic risk scores generated from unconditioned GWAS summary statistics (Figure 1, bottom row). Individuals in the highest 20% of conditioned IA polygenic risk had a pooled hazard ratio of 1.96 (95% CI 1.11-3.46, p = 0.020, heterogeneity p=0.609) compared to the lowest 20% of polygenic risk. We did not find an association between hypertension PRS or smoking intensity PRS and risk of SAH or IA (Kaplan-Meier cumulative incidence curves for each cohort shown in Supplementary Figures 1-3). We found no difference in cumulative incidence of SAH or IA between *PKD1* or *PKD2* monogenic variant carriers, nor between carriers of *PKD1* truncating or non-truncating variant carriers (Supplementary Figures 4-5).

**Figure 1.**
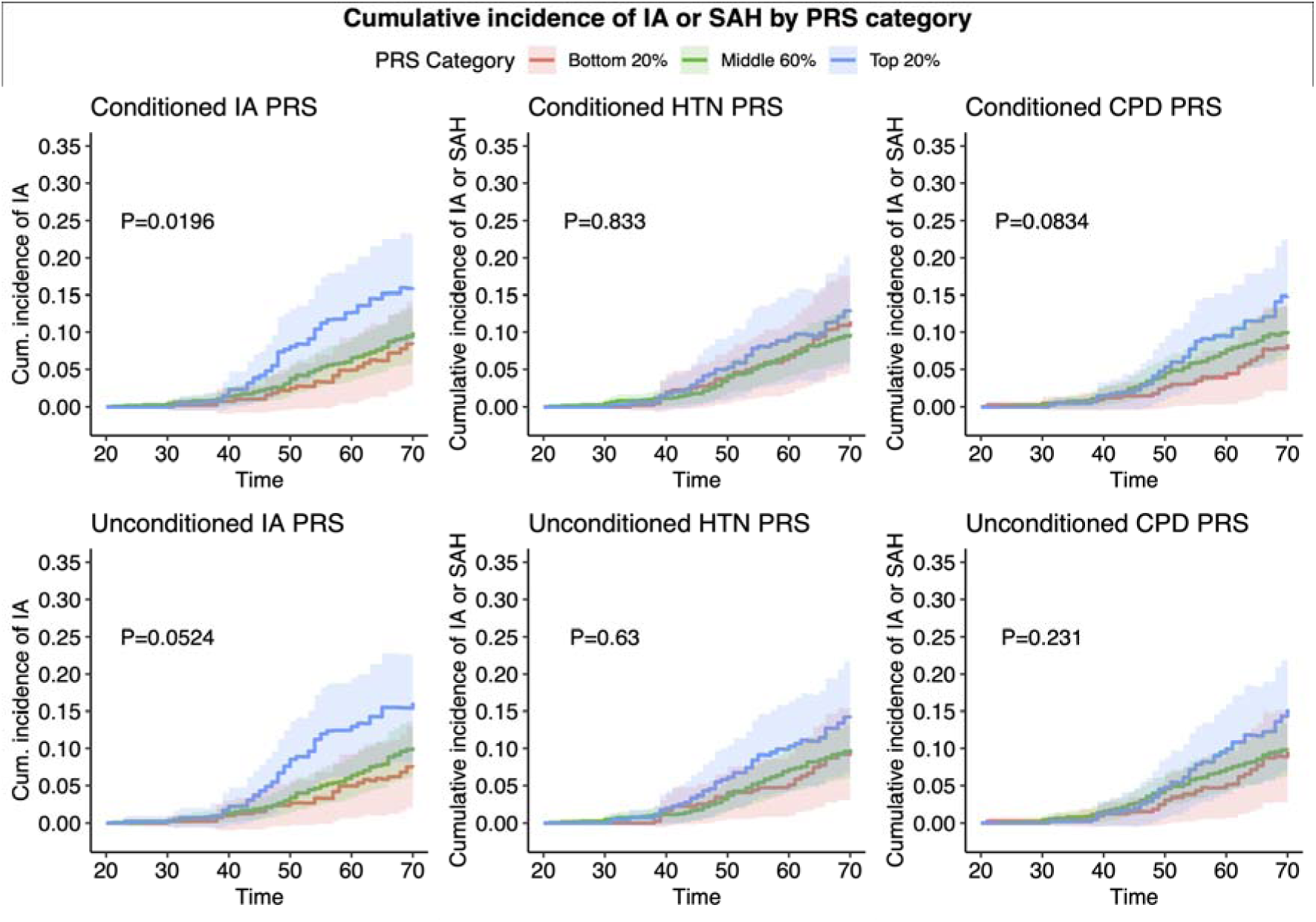
Cumulative incidence of intracranial aneurysm or subarachnoid haemorrhage in ADPKD. Pooled cumulative incidence curves across FinnGen, GEL, and IKGP cohorts. Cumulative incidence of IA by intracranial aneurysm (IA) PRS, hypertension (HTN) PRS, and smoking intensity (cigarettes per day, CPD) PRS. The top row represents polygenic scores calculated on mtCOJO conditioned GWAS summary statistics, while the bottom row represents polygenic scores calculated from original unconditioned GWAS summary statistics. P-value represents meta-analysis across cohorts comparing top 20% of polygenic score risk to bottom 20% of polygenic score risk.

### Smoking, female sex, and polygenic risk as risk factors in univariate analysis

We found in an inverse-variance weighted meta-analysis of univariate Cox regression that smoking and female sex were significant clinical risk factors for IA or SAH in individuals with ADPKD, with a pooled hazard ratio for smoking status of 2.06 (95% CI 1.43-2.96, p=9.49 x10^−5^) and a pooled hazard ratio for female sex of 2.21 (95% CI 1.50-3.25, p=6.52×10^−5^) (Figure 2). We also found that diagnosis of Type 1 or Type 2 diabetes was associated with a reduced hazard of IA in individuals with ADPKD (HR 0.39, 95% CI 0.22-0.70, p=1.37×10^−3^) compared to those that did not have diabetes. We found in univariate analysis that both the conditioned and unconditioned IA polygenic score, when assessed as a continuous variable, significantly predicted higher risk of IA, with the conditioned PRS giving a pooled hazard ratio of 1.30 (95% CI 1.09-1.57, p=4.66×10^−3^) and unconditioned PRS giving a pooled hazard ratio of 1.29 (95% CI 1.07-1.54, p=6.21 x10^−3^), representing a higher risk with 1 standard deviation increase in polygenic score. We did not find evidence of an increased risk of IA or SAH in individuals with *PKD1* monogenic variants compared to *PKD2* carriers or individuals with no causal variant found, nor an increased risk in *PKD1* truncating variant carriers compared to *PKD1* non-truncating variant carriers in the pooled meta-analysis. Heterogeneity among cohort-specific Cox regression results was significant for smoking (p<0.001) and dyslipidaemia (hypercholesterolemia) (p=0.017) but was not detected among other variables. Full Cox regression summary statistics for the data represented in Figure 2 are shown in Supplementary Table1.

**Figure 2.**
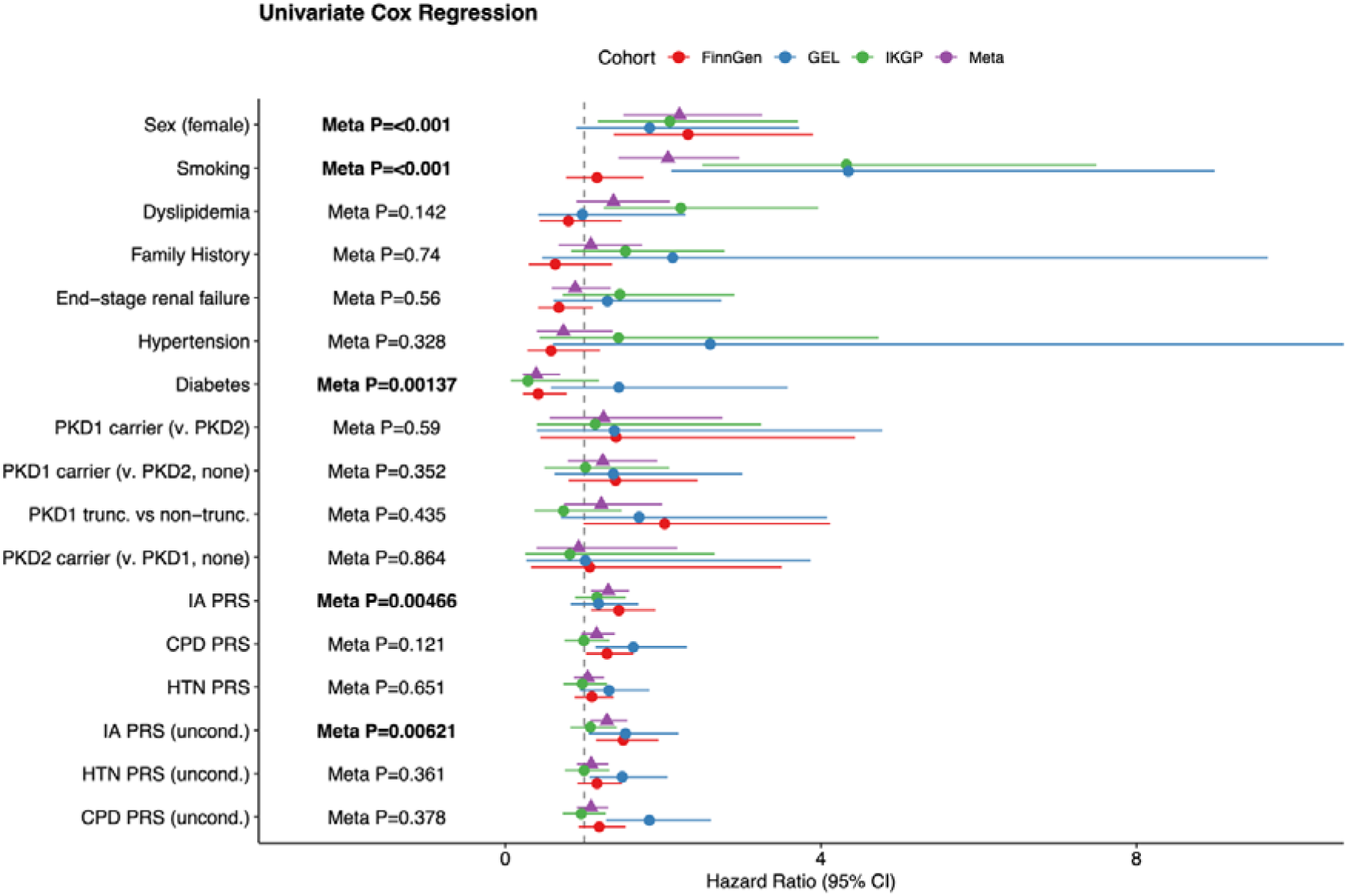
Univariate analysis of clinical and genetic risk factors for IA. Cox proportional hazards model for each risk factor. IA = Intracranial Aneurysm, HTN = Hypertension, CPD = smoking intensity, cigarettes per day. uncond. = polygenic score calculated from original GWAS summary statistics, cond. = polygenic score calculated from mtCOJO-conditioned GWAS summary statistics. Shapes represent pooled (meta-analysis, triangle) or original (circles) hazard ratio from Cox univariate regression analysis with first 10 PCs of genetic variation as covariates, while bars represent 95% confidence intervals. P values represent inverse-variance weighted meta-analysis of p values from each cohort. Full p-values and hazard ratios for each cohort are provided in Supplementary Table 1.

### Smoking, female sex, and polygenic risk remain in multivariate analysis

To assess the joint role of clinical factors, monogenic variants, and polygenic scores, we performed multivariate Cox regression analysis with either conditioned polygenic risk scores (Figure 3A) or unconditioned polygenic risk scores (Figure 3B). We found that even after accounting for clinical variables (sex, smoking status, dyslipidaemia, family history, end-stage renal failure, hypertension, and diabetes status), both conditioned and unconditioned IA polygenic scores were significantly associated with IA or SAH, with conditioned IA PRS giving a pooled hazard ratio of 1.26 (95% CI 1.03-1.55, p=0.025) and unconditioned IA PRS giving a pooled HR of 1.28 (95% CI 1.04-1.59, p=0.019). Smoking and female sex remained significant risk factors for IA or SAH in the multivariate model (smoking HR 2.85, 95% CI 1.88-4.31, p=7.40×10^−7^, female sex HR 1.98, 95% CI 1.28-3.07, p=2.09×10^−3^, conditioned PRS model), and Type 1 or Type 2 diabetes remained a significant protective factor against IA or SAH (HR 0.35, 95% CI 0.17-0.69, p=2.54×10^−3^, conditioned PRS model). A nominally significant effect of hypertension was detected in the unconditioned PRS model (p=0.04), but this is likely due to over-adjustment in the model as variance inflation factors were all below 3 (Supplementary Table 2). Significant heterogeneity among cohort-specific Cox regression results was detected in the effect of smoking (p<0.001), but not in any other variable in the model. Full Cox regression model results for Figure 3 are given in Supplementary Table 3.

**Figure 3.**
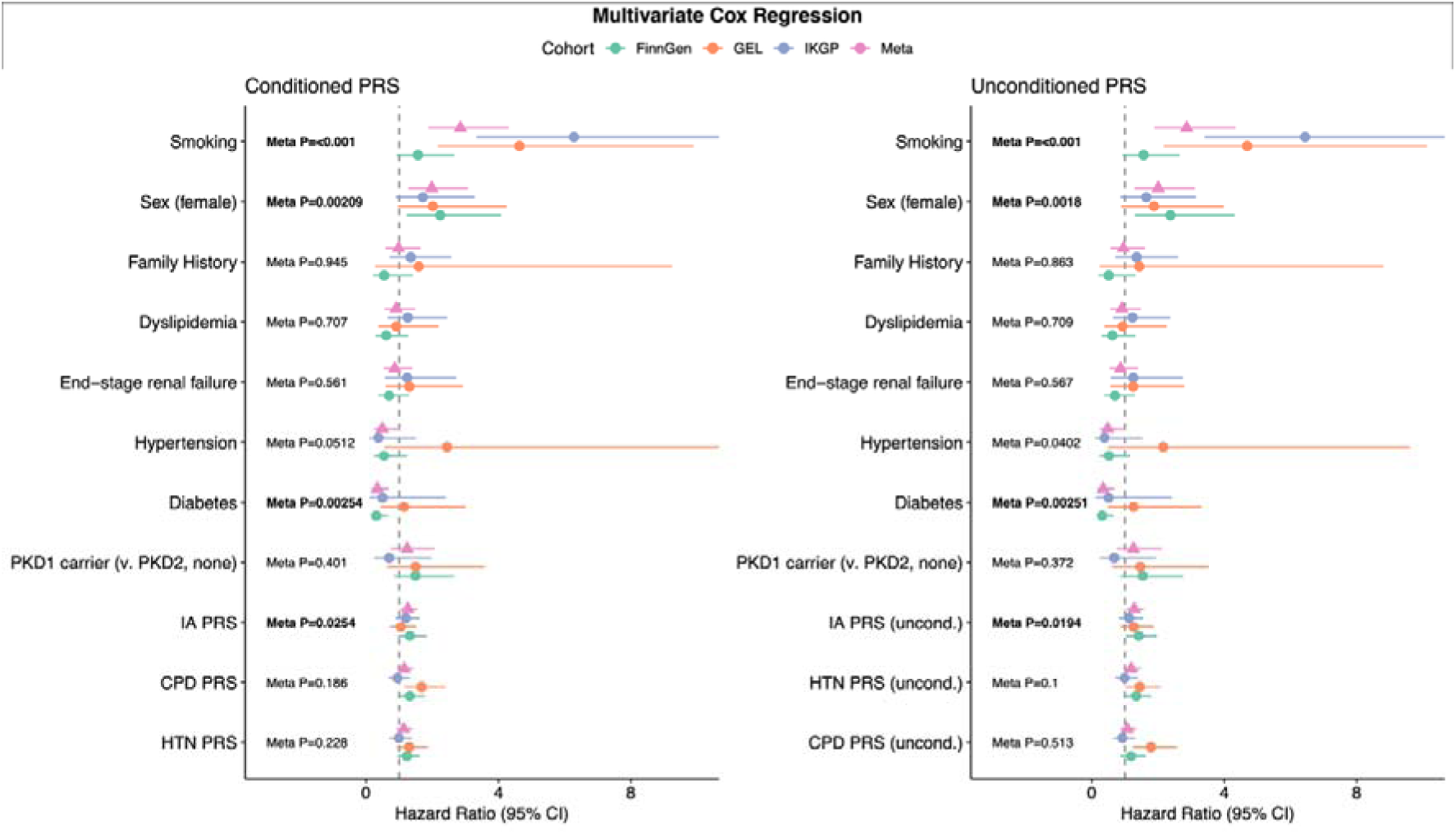
Multivariate analysis of clinical and genetic risk factors for IA. Cox proportional hazards model for risk factors in a joint multivariate model, additionally adjusted by first ten principal components of genetic information. A) Multivariate model including conditioned polygenic risk scores, B) multivariate model including unconditioned polygenic risk scores. IA = Intracranial Aneurysm, HTN = Hypertension, CPD = smoking intensity, cigarettes per day. uncond. = polygenic score calculated from unconditioned GWAS summary statistics, conditioned = polygenic score calculated from mtCOJO-conditioned GWAS summary statistics. Full p-values and hazard ratios for each cohort are provided in Supplementary Table 3.

### Assessment of clinical significance

Model discrimination via concordance index (C-index) varied markedly by cohort (Figure 4A). In FinnGen, the base model containing clinical variables (smoking status, sex, family history of IA or SAH, end-stage renal failure, dyslipidaemia, hypertension, diabetes) had a C-index of 0.75 (95% CI 0.69-0.81), while the model including baseline clinical variables and polygenic risk scores had a C-index of 0.77 (95% CI 0.72-0.83) and the nested model containing baseline clinical variables, monogenic variant status, and IA polygenic risk score had a C-index of 0.78 (95% CI 0.73-0.84). Comparing the base clinical model to the polygenic model with the likelihood ratio test, the polygenic model significantly better predicted IA or SAH (p=3.61×10^−3^). We found a similar effect in GEL, where the model including polygenic scores significantly better predicted IA or SAH than base clinical variables alone (p=8.84×10^−3^) (Figure 4A, comparison in all cohorts in Supplementary Table 4). In contrast, the C-index values for IKGP were similar for all models (Figure 4A), with no significant difference between models on the likelihood ratio test (Supplementary Table 4). Consequently, meta-analysis of C-index among cohorts gave wide error estimates, with significant heterogeneity among cohorts (p=1.61×10^−6^ – 7.82×10^−4^, Supplementary Table 5).

**Figure 4.**
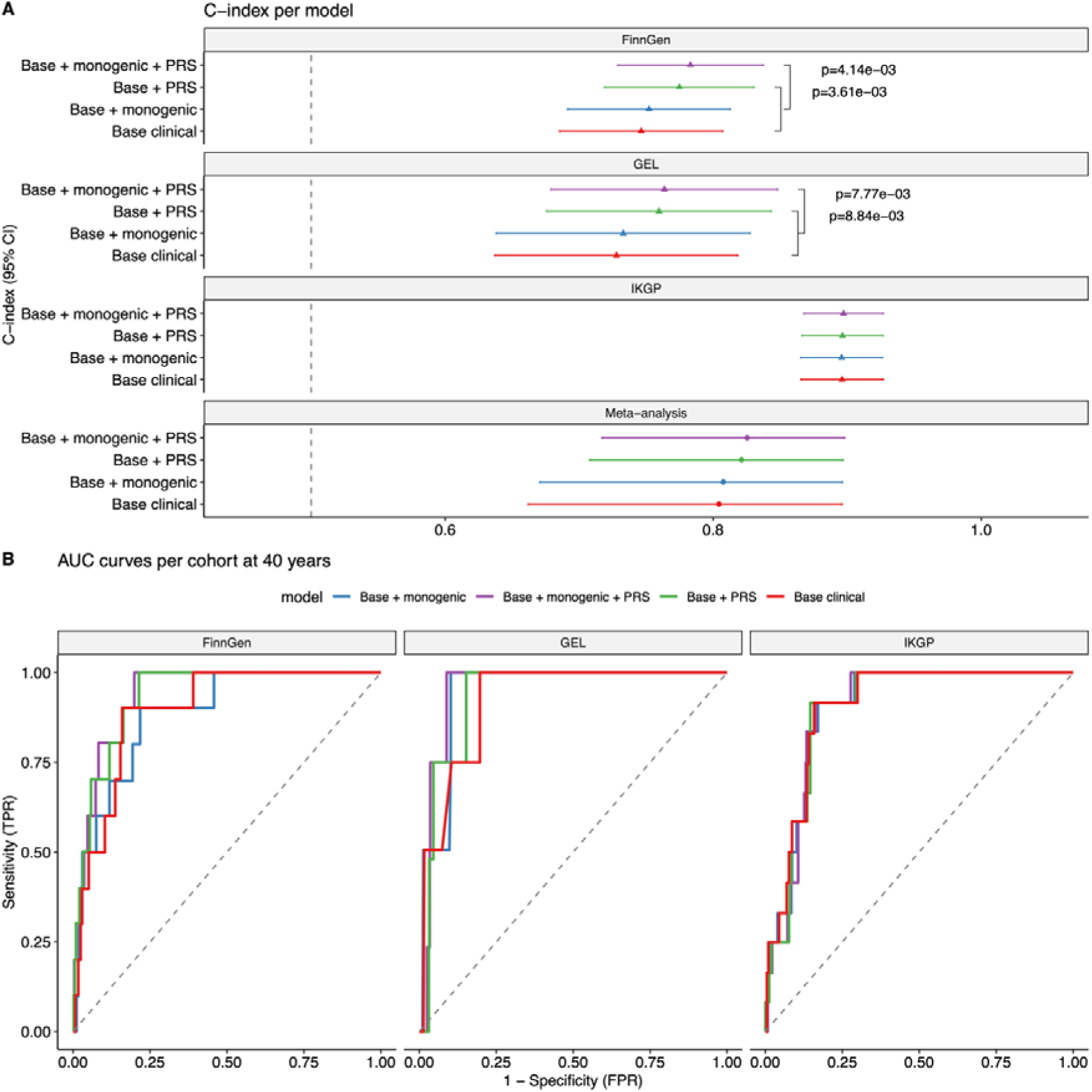
Clinical utility of additional genetic information in predicting IA. A) C-index comparing four nested models: base clinical model consisting of age, sex, smoking status, family history, end-stage renal failure, dyslipidaemia, hypertension, and diabetes status, base clinical model plus IA, CPD, and HTN conditioned polygenic scores, base clinical model plus PKD1, PKD2 or no rare variant found, and base clinical model plus rare variant status and PRS. P-values represents analysis of variance (ANOVA) comparing two nested models, e.g. the likelihood ratio test. B) time-dependent ROC curves showing sensitivity (true positive rate, TPR) vs specificity (false positive rate, FPR) in all three models, for each cohort.

This heterogeneity also reflects in the visualization of meta-analysed time-dependent ROC curves (Figure 4B), which show poor discrimination among models in IKGP but better discrimination in FinnGen and GEL. Time-dependent ROC curves at age 40, 50, 60, and 70 are shown for each cohort and as a pooled meta-analysis in Supplementary Figures 6-9.

### Mendelian randomisation

We performed a multiple instrumental variable Mendelian Randomisation analysis using SNPs associated with hypertension in the FinnGen cohort (significant with a p-value of <10^−7^) as the exposure data and the summary statistics from Bakker *et al.*^27^ as the outcome data, according to the risk of IAs and SAH, respectively. The analysis showed that there is a significant association of hypertension with the risk of both IAs and SAH (Supplementary Data 1-2; Supplementary Figure 10-11). Further sensitivity analyses with MR-Egger and Weighted median methods, which allow violation of some MR assumptions, showed that the association remained significant (Supplementary Figures 10-11). This association with hypertension emphasizes the role of hypertension in IA and SAH in a general population.

## Discussion

This multi-cohort study provides evidence that polygenic risk, smoking, and female sex are significant risk factors for IA or SAH in patients with polycystic kidney disease. Individuals with the highest 20% polygenic risk for IA had a pooled hazard ratio of 1.96 (95% CI 1.11-3.46) for IA development or rupture compared to the cohort with lowest 20% PRS for IA in univariate analysis, and in multivariate analysis accounting for other clinical factors the IA PRS remained significant with a hazard ratio of 1.26 (95% CI 1.03-1.55). Smoking and female sex had largest effect on IA or SAH risk in ADPKD patients, with hazard ratios of 2.85 (95%CI 1.88-4.31) and 1.98 (95% CI 1.28-3.07), respectively, in multivariate analysis. Neither monogenic variant status nor hypertension were found to associate with increased IA or SAH risk, although the latter is likely due to high prevalence of hypertension in this patient population. Together, these findings suggest that both genetic predisposition and modifiable lifestyle factors shape IA or SAH risk in ADPKD.

Effect size estimates for smoking varied across populations, likely reflecting differences in how smoking history was ascertained, but the association remained strong and consistent in direction, reinforcing the central importance of smoking cessation counselling. The magnitude of the smoking effect is in line with previous studies in general populations^32–34^, which identify smoking as the strongest modifiable risk factor and a causative factor for aneurysm formation and rupture^35,36^. Our study found, however, that genetic predisposition to cigarette smoking intensity as assessed by polygenic score was not associated with IA or SAH risk. Although the Bakker *et al.* GWAS identified a significant genetic contribution to IA from smoking intensity and hypertension liability^27^, genetic predisposition to smoking intensity per se may not increase risk for IA unless smoking is first initiated and is therefore not as useful a predictor of IA risk than smoking status itself.

Our observed higher risk of IA or SAH with female sex is consistent with prior epidemiological data showing higher IA prevalence in women, particularly after the menopause^37–39^. Hormonal factors, including the effects of oestrogen on vascular wall remodelling and collagen synthesis, have been proposed as mechanisms underlying this sex-specific vulnerability^40,41^, although the mechanism is incompletely understood.

Type 1 or Type 2 diabetes was a significant protective factor for IA (HR 0.35, 95% CI 0.17-0.69), consistent with prior observational and genetic studies in a general population, including a nationwide Korean cohort^42^and a Mendelian randomisation analysis using the same IA GWAS that informed our polygenic score^43^. Several mechanisms have been proposed, including hyperglycaemia-induced arterial wall stiffening through accumulation of advanced glycation end-products and altered vascular remodelling, which together may counteract the haemodynamic and structural processes that drive aneurysm formation. Tighter blood pressure control in this group is a likely additional contributor: patients with diabetes are commonly managed to lower blood pressure targets and are frequently treated with ACE inhibitors or angiotensin receptor blockers for renal and cardiac indications, and this residual blood pressure effect would not be fully captured by adjusting for a binary hypertension diagnosis. Specific glucose-lowering therapies may also play a role, with metformin^44^ and, more recently, GLP-1 receptor agonists^45^ both associated with reduced SAH risk in diabetic patients with IAs.

This protective association has recently been reinforced by the largest systematic review and meta-analysis to date, which confirmed a reduced risk of aneurysmal SAH (aSAH) in diabetic individuals without sex-specific differences^46^. However, a separate meta-analysis examining rupture risk specifically in patients with known IAs^47^ did not find an association, suggesting that the protective effect of diabetes may relate more to aneurysm formation and the development of a SAH rather than at the point of rupture once an IA is established. Our results extend this observation to an ADPKD population, though larger studies will be needed to confirm whether the effect is consistent across monogenic genotypes and treatment patterns.

Hypertension affects approximately 75-80% of ADPKD patients and typically develops earlier than in the general population^48^. Neither hypertension nor genetic predisposition to hypertension measured via polygenic score was associated with IA or SAH risk in the present analysis. The prevalence of hypertension was very high across cohorts, consistent with previous reports in ADPKD^48^, raising the possibility that hypertension is so prevalent in this group that it does not discriminate between individuals with and without IA in the way that clinical factors with lower prevalence such as smoking, diabetes, and sex are able to. Our multivariable Mendelian randomisation analysis from GWAS summary statistics derived from a general population emphasized that hypertension plays an important role in IA or SAH, but the contrast with the lack of an observed association between hypertension and IA in ADPKD likely reflects the near universal prevalence of hypertension in this group of individuals rather than absence of a true causal effect.

The role of *PKD1* and *PKD2* in the elevated IA risk seen in ADPKD remains incompletely characterised, and whether risk differs meaningfully between the two genes has been debated^49^. The largest cross-sectional study to date, the Genkyst cohort, reported a higher IA diagnosis rate in *PKD1* compared with *PKD2* carriers and identified *PKD1* monogenic genotype as an independent risk factor in multivariable analysis, though neither variant type nor position within *PKD1* modified this risk^24^. A smaller Japanese single-centre study subsequently reported earlier IA onset specifically in carriers of PKD1 truncating variants, suggesting that variant class may also influence risk^25^. More recently, a population-level analysis of UK Biobank exome data demonstrated that, even outside of a confirmed ADPKD diagnosis, rare damaging PKD1 variants are independently associated with IA risk in the general population, with only a nominal signal for PKD2^26^. In our study, we found no association between monogenic variants in *PKD1* or *PKD2* and increased risk of IA or SAH. This is most likely due to the apparent effect of *PKD1* variant being absorbed by clinical risk factors, suggesting that monogenic contribution to IA in ADPKD does not act in isolation but on a background of broader genetic and environmental susceptibility, although heterogeneity in the definitions of *PKD1* and *PKD2* causal variants may also play a role. Overall, our study jointly modelled rare monogenic variants, polygenic background, and clinical risk factors in the largest multinational study to date, and the apparent attenuation of the monogenic signal in this combined framework warrants confirmation in larger cohorts.

In the present analysis, family history did not show a significant association with IA or SAH risk, contrasting with some previous studies^50^ but in keeping with others^13,51^. This lack of association between IA and SAH and family history was irrespective of ascertainment of family history, which differed markedly among cohorts–ICD codes in Genomics England, clinical records and pedigree confirmation in IKGP, and genetically-inferred ancestry and EHR-derived diagnoses in FinnGen. Family history is often considered a proxy and inexpensive alternative to genetic risk profiling^7^, yet family history can be both inaccurately reported and represents both shared environmental risk factors and inherited genetic risk, the latter of which can vary within individuals within a family depending on the variants they inherit^23^. Our finding of a significant association between IA or SAH and IA polygenic score but not family history suggests that direct assessment of genetic liability to IA in ADPKD patients is a more effective estimate of risk, in line with previous studies that have shown PRS to be complementary to family history^22^, and that current screening guidelines for IA in patients with ADPKD may not effectively capture individuals at the highest risk.

However, while our study demonstrated that polygenic predisposition for IA as assessed by IA polygenic score is significantly associated with IA risk in patients with polycystic kidney disease, evidence for the clinical utility of IA PRS is mixed. While models including polygenic risk scores significantly improved discrimination of individuals at risk of IA or SAH in FinnGen and GEL over clinical risk factors alone, the addition of genetic information in IKGP did not significantly improve discrimination. This may reflect more thorough ascertainment of IA risk through clinical variables in IKGP, where information is gathered via structured clinical consultation. In FinnGen and GEL, where data derive solely from electronic health records and ascertainment may therefore be incomplete or biased, genetic risk factors meaningfully improved discrimination. This suggests that incorporating genetic information could help stratify IA risk among ADPKD patients in settings where clinical assessment is not systematic– for example, in large hospital systems where automated, EHR-linked genetic risk scoring could flag high-risk patients who may otherwise go unidentified. Whether such an approach is economically justified relative to the costs of care or mortality associated with an undetected, ruptured SAH remains to be established.

Several limitations warrant consideration. First, the retrospective nature of the analysis and potential ascertainment bias in the cohorts may affect the generalisability of findings. Second, the polygenic risk scores were derived from populations of primarily European ancestry, potentially limiting their applicability to other ethnic groups. Recent research has highlighted the importance of multi-ancestry polygenic risk score development to address health disparities and ensure equitable implementation of genetic risk prediction^28,52^. Third, we lacked detailed information on IA characteristics such as size, location, and morphology, which could provide additional insights into genetic determinants of aneurysm phenotypes and were unable to analyse IA and SAH as separate outcomes due to the limited number of events. Furthermore, not all patients underwent screening for IAs, there were significant differences in this practice between cohorts and was undoubtedly influenced by individual clinician practices. Finally, information on some clinical risk factors had markedly different ascertainment among cohorts, notably smoking status and family history of IA. However, despite limitations this study remains the largest multinational study to date, and among the first to jointly assess the role of clinical risk factors, rare monogenic variants, and polygenic background.

### Conclusions

This study demonstrates that smoking status, female sex, and polygenic risk for IA are important risk factors for IA or SAH risk in ADPKD patients. Hypertension was not found to be associated with IA or SAH, likely due to high prevalence in this patient population. These findings suggest that current screening guidelines targeting family history of IA or SAH may not capture the highest individuals at risk and suggest that personalised screening strategies utilising genetic risk may improve risk stratification in ADPKD, although clinical utility of polygenic risk scores in general remain to be verified.

## Methods

### Cohorts

We conducted a collaborative multi-cohort study using data from three independent populations: Ireland (Irish Kidney Gene Project, IKGP)^53^, Finland (FinnGen)^54^ and the United Kingdom (Genomics England Limited, GEL)^55^. Briefly, IKGP is a national research initiative with kidney disease patients recruited from nephrology services across Ireland, combining biological samples, tissue samples, and omics data with detailed clinical data. FinnGen is a public-private research initiative composed of disease-focused legacy cohorts and prospectively recruited samples from clinical contacts throughout Finland, with access to genomics data and rich longitudinal electronic health record data. Genomics England’s 100,000 Genomes Project is a disease-focused program focused on whole-genome sequencing of patients with rare diseases, cancer, and their relatives, combined with electronic health record data. Detailed information on each cohort, including ethical statements, is provided in the supplementary methods.

### Clinical Data Collection

Clinical variables were assessed for each cohort, including identifying cases of polycystic kidney disease, ruptured or unruptured IAs, and smoking status, sex, family history of IA or SAH, dyslipidaemia, end-stage kidney disease, hypertension, and diabetes. Clinical data collection varied depending on the cohort and available data for diagnosis. Briefly, clinical data for FinnGen and GEL were extracted from pseudonymized electronic health record data composed only of structured, coded data, while in IKGP clinical data was derived from full electronic patient records with clinical notes available. In FinnGen clinical variables were derived from clinically curated endpoints drawing from multiple data registers (inpatient, outpatient, and cause of death registers, among others) with ICD codes, prescription data, and hospital procedure codes. Additional strict criteria applied for IA/SAH case identification was applied in FinnGen. In GEL, predefined coding frameworks were used to extract diagnosis data from Hospital Episode Statistics records, namely relevant ICD-10 and SNOMED codes from inpatient, outpatient, emergency and intensive care records (Supplementary Data 3). In IKGP, clinical variables, family history and imaging findings were extracted directly from electronic healthcare records (E-Med, Progeny and NIMIS) by manual chart and imaging review, with IA and SAH cases confirmed against the corresponding cerebral imaging and radiology reports. Of note, FinnGen included SAH cases from the Finnish cause of death register, while deaths due to SAH occurring outside the hospital were not captured in the IKGP and GEL cohorts. To improve sample size for multivariate analyses, smoking status was imputed for individuals missing direct smoking information in IKGP and FinnGen cohorts. Detailed information for phenotype definitions for each cohort, including smoking imputation methods, is presented in the supplementary methods.

### Imaging confirmation of IA/SAH

Radiology imaging reports (MRA, CT angiogram, DSA and or direct cerebral angiogram) were reviewed for IKGP cases where available to determine which scans were performed for screening purposes and which were performed for cause to evaluate potential SAH. In FinnGen, imaging status was determined with NOMESCO procedure codes, while in GEL imaging was defined with SNOMED CT codes.

Further detailed information on imaging confirmation is provided in the supplementary methods.

### Monogenic variant evaluation

Methods for evaluation and identification of rare, monogenic genetic variants associated with polycystic kidney disease are described in detail for each cohort in the supplementary methods. Briefly, in IKGP monogenic variants were identified with a combination of next-generation sequencing strategies, including whole-exome sequencing and targeted kidney gene panels covering known cystic and chronic kidney disease genes. In FinnGen, monogenic variants were identified with whole-exome sequencing by applying strict quality control and identifying either variants previously associated with polycystic kidney disease in ClinVar^56^ or novel variants with in-silico predicted damaging effects. In GEL, monogenic variants were identified via whole –genome sequencing, also applying strict quality control and identifying in-silico predicted damaging variants.

### Polygenic Risk Score Calculation

Polygenic risk scores were calculated for three traits: intracranial aneurysms (IA)^27^, hypertension (HTN)^57^, and cigarettes per day (CPD)^29^. Detailed information on polygenic score calculation and quality control of genotyping data used for polygenic scores is described in detail in the supplementary methods. Polygenic scores were calculated in two ways: with unconditioned and conditioned GWAS summary statistics. For polygenic risk scores based on unconditioned GWAS summary statistics, we followed typical PRS calculation methods. For polygenic risk scores based on conditioned GWAS summary statistics, we first conditioned the effect sizes of each phenotype’s genetic variants on the remaining two phenotypes with mtCOJO^31,58^. In other words, the conditioned effect sizes for SNPs associated with IA from Bakker et al^27^ reflect the effect on IA after accounting for their effect on smoking intensity (cigarettes per day)^29^ and on hypertension^59^, as assessed by genome-wide association studies for these traits. After conditioning each phenotype’s GWAS summary statistic on the remaining two phenotypes, we followed an identical protocol as above to calculate polygenic scores.

### Cumulative incidence curves

Cumulative incidence curves were estimated using the Kaplan-Meier method (survfit, R survival package) as 1 – S(t) or one minus the survival function for time from birth to age of record of diagnosis of IA or SAH. For survival analysis, participants were stratified into bottom 20%, middle 60%, and top 20% of polygenic risk for IA, hypertension, and smoking intensity (cigarettes per day), for both conditioned and unconditioned polygenic scores. Time-to-event analysis was conducted with appropriate censoring for loss to follow-up. Survival analysis was run independently within each cohort, with polygenic risk stratification determined independently within each cohort to account for potential population-specific genetic architecture differences.

All three cohorts were then combined to create a weighted survival curve, where pooled survival was calculated as a weighted mean of survival at each time point with weighting equivalent to the risk set in each cohort at that time point and total risk set was the sum of all individuals in all cohorts at risk at that time point. Detailed information on the weighted survival curves is given in the supplementary methods.

Survival curves were also estimated with the same method for monogenic variant carrier status, stratifying on *PKD1*, *PKD2*, or no variant found (removing individuals from the analysis carrying rare variants other than in *PKD1* or *PKD2* and combining truncating and non-truncating variants for each gene). Additional survival curves were estimated for *PKD1* truncating and non-truncating variants. Survival curves were estimated independently in each cohort, and pooled survival curves were calculated with the method described in the supplementary methods.

### Univariate and multivariate and Cox regression analysis

Univariate Cox proportional hazards models were fit for each clinical variable of interest (smoking status, sex, family history of IA or SAH, dyslipidaemia, end-stage kidney disease or ESKD, hypertension, diabetes, defined in detail in Supplementary Methods), conditioned and unconditioned polygenic scores as continuous variables (IA, hypertension, and cigarettes per day, i.e. smoking intensity), or rare variant status (*PKD1* carrier vs *PKD2* carrier vs no variant found, *PKD1* vs *PKD2* rare variant carrier, and *PKD1* truncating vs non-truncating variant). Smoking status in FinnGen was missing for 235 (28.5%) individuals and was handled using M=25 imputations, as described in the supplementary methods; the resulting datasets were stacked into a single analytic dataset in which each individual contributed 25 rows, each weighted by 1/25, and Cox proportional hazards models were fitted with robust sandwich standard errors clustered on individual identifier to account for the non-independence of imputed observations. In IKGP and GEL, smoking status was modelled as a three-level factor, with reference level ‘never smoker’ and contrast levels ‘unknown smoking status’ and ‘ever smoker’.

For IKGP and GEL family history of IA and SAH was modelled as a three-level factor with ‘unknown history’ as a contrast level (in FinnGen, with genetically determined relationships and family history from ICD codes, family history was determined as a two-level factor). Each univariate Cox proportional hazards model was fit with coxph (R survival package) with time-to-event defined via Surv(time, IA or SAH), with covariates as first 10 principal components of genetic information to account for population stratification.

Two multivariate Cox proportional hazards were fit to assess the joint effect of clinical risk factors, monogenic variants, and polygenic risk. The first model included clinical risk variables specified above, as well as monogenic variants carrier status (*PKD1* carrier vs *PKD2* carrier vs no variant found), unconditioned polygenic scores as continuous variables, and first 10 PCs. The second model included the same clinical risk variables, monogenic variant carrier status, and first 10 PCs, as well as conditioned polygenic scores.

### Assessing clinical significance

Model discrimination was assessed using the concordance index (C-index), the survival analysis equivalent of the area under the ROC curve (AUC). The C-index represents the probability that, for a randomly selected pair of patients where one experienced the event first, the model assigned that patient a higher predicted risk. C-indices were estimated from four nested Cox models: clinical variables only, clinical variables and polygenic risk scores, clinical variables and monogenic variant status, and clinical variables, monogenic variant status, and conditioned intracranial aneurysm polygenic score. C-indices were estimated via concordance () (R survival package), with 95% confidence intervals computed from the variance of the concordance statistic. C-indices were estimated individually for each cohort, and meta-analysed across cohorts with random-effect inverse-variance-weighted meta-analysis. In FinnGen, c-index was derived directly from the stacked weighted Cox models, with robust standard errors accounting for clustering by individual.

Nested Cox proportional hazards models were compared using the likelihood ratio test (LRT) (anova), R survival package). Each successive model was compared against the prior model to assess whether the addition of covariates produced a statistically significant improvement in model fit. In FinnGen, LRT was performed by fitting nested Cox models separately within each of the 25 datasets (reflecting 25 smoking imputation iterations) without weights or clustering, and the resulting chi-squared statistics were pooled by averaging across imputations, with a single p-value derived from the pooled statistic against a chi-squared distribution with the appropriate degrees of freedom.

To visualize the sensitivity and specificity of the models, time-dependent receiver operating characteristic (ROC) curves were constructed at prespecified time horizons (AUC at 40 years, 50 years, 60 years, and 70 years of age for using the Blanche et al. (2013) estimator (timeROC(), R timeROC package). Unlike standard ROC analysis, this approach accounts for censoring by defining case/control status as time-dependent, such that discrimination is evaluated at a fixed time horizon t.

The area under the time-dependent ROC curve (AUC(t)) was estimated at each horizon with a 95% confidence interval derived from the influence function (iid decomposition). For FinnGen, linear predictors from the stacked Cox models were averaged across the 25 imputations to yield a single risk score per individual prior to ROC estimation. To pool curves across cohorts, each cohort’s ROC curve was interpolated onto a common false positive rate grid (0-1 step 0.005) using linear interpolation. At each grid point, a pooled TPR was computed as an inverse-variance weighted mean across cohorts, weighted by the reciprocal of the squared AUC standard error, with 95% confidence intervals derived from the pooled standard error. AUC values were pooled separately on the logit scale using an inverse-variance weighted mean, with back-transformation to the probability scale for reporting.

## Data Availability

All data produced in the present study are available upon reasonable request to the authors

## Supplementary Information

### List of supplementary files

**Supplementary Data 1 and 2**

Multiple instrumental variable Mendelian Randomization per-snp and aggregated results for hypertension exposures on ruptured and unruptured intracranial aneurysms.

**Supplementary Data 3:** codes used to define phenotypes in the Genomics England cohort

### Supplementary Figures

1. **Supplementary Figure 1** Intracranial aneurysm polygenic risk stratifying IA or SAH in each cohort
2. **Supplementary Figure 2** Hypertension polygenic risk stratifying IA or SAH in each cohort
3. **Supplementary Figure 3** Smoking intensity polygenic risk stratifying IA or SAH in each cohort
4. **Supplementary Figure 4** Cumulative incidence of IA or SAH by monogenic variant status, meta-analysis
5. **Supplementary Figure 5** Cumulative incidence of IA or SAH by monogenic variant status, per cohort
6. **Supplementary Figure 6** FinnGen time-dependent ROC curves
7. **Supplementary Figure 7** GEL time-dependent ROC curves
8. **Supplementary Figure 8** IKGP time-dependent ROC curves
9. **Supplementary Figure 9** Meta-analysed time-dependent ROC curves
10. **Supplementary Figure 10** Mendelian Randomization of hypertension on unruptured IA
11. **Supplementary Figure 11** Mendelian Randomization of hypertension on SAH

### Supplementary Tables

1. **Supplementary Table 1** Results of univariate Cox regression models
2. **Supplementary Table 2** Variance inflation factors for multivariate Cox regression models
3. **Supplementary Table 3** Results of multivariate Cox regression models
4. **Supplementary Table 4** Comparison of models for all cohorts
5. **Supplementary Table 5** Meta-analysis of c-index across cohorts

## Supplementary Methods

### Cohorts

The Irish Kidney Gene Project (IKGP)^53,60^ is a national research initiative established in 2014 to characterise the genetic basis of kidney disease in Ireland. The project is led jointly by Beaumont Hospital and the Royal College of Surgeons in Ireland (RCSI), with patients recruited from nephrology services across the country. To date, more than 1,800 patients with suspected inherited kidney disease have been enrolled, with a molecular diagnosis identified in approximately half. The IKGP combines a national biobank of DNA, blood, urine, and kidney tissue samples with detailed clinical data, and operates a dedicated genetic kidney disease clinic offering diagnosis and counselling. Through international collaborations and the use of next-generation sequencing, the project aims to discover novel disease-causing genes, refine prognostication, and ultimately translate genetic findings into new therapies for patients with kidney disease.

FinnGen^54^ is a public-private research initiative established in 2017 that integrates genomic data with digital health records from approximately 500,000 Finnish individuals. The project brings together Finnish biobanks, universities, university hospitals, and international pharmaceutical partners under a pre-competitive framework, with coordination supported by the Finnish biobank cooperative FINBB. Its overarching goal is to generate clinically and therapeutically meaningful insights into human disease at a population scale.

Genomics England’s 100,000 Genomes Project is a large disease-focused sequencing programme generating whole-genome data from NHS patients with rare diseases, cancer, and their relatives^55^. Its strengths include uniform processing through a shared pipeline and the availability of population-matched controls without the phenotype of interest, enabling robust control of technical artefacts, allele frequency, and variant burden. Recruitment to the 100,000 Genomes Project was conducted through 13 NHS Genomic Medicine Centres, with phenotypic data captured using hierarchically structured HPO terms to enable systematic computational analysis. Using Genomics England dataset v15, comprising over 90,000 participants with linked genomic and clinical data, we applied quality control, relatedness filtering, and ancestry matching, yielding 899 European-ancestry cases for analysis^61^

### Ethical Considerations

Ethical approval and data sharing agreements were obtained from the relevant institutional review boards and committees for each of the cohorts.

#### Ethical statement IKGP

This study was conducted as part of the Irish Kidney Gene Project (IKGP). Ethical approval was granted by the Beaumont Hospital Ethics (Medical Research) Committee (REC reference 19/28). The study was performed in accordance with the Declaration of Helsinki and applicable Irish and European Union legislation, including the General Data Protection Regulation (GDPR). All participants provided written informed consent for clinical data collection, genetic analysis, biobanking of biological samples, and the sharing of de-identified data with approved research collaborators.

#### Ethical Statement FinnGen

Study subjects in FinnGen provided informed consent for biobank research, based on the Finnish Biobank Act. Alternatively, separate research cohorts, collected prior the Finnish Biobank Act came into effect (in September 2013) and start of FinnGen (August 2017), were collected based on study-specific consents and later transferred to the Finnish biobanks after approval by Fimea (Finnish Medicines Agency), the National Supervisory Authority for Welfare and Health. Recruitment protocols followed the biobank protocols approved by Fimea. The Coordinating Ethics Committee of the Hospital District of Helsinki and Uusimaa (HUS) statement number for the FinnGen study is Nr HUS/990/2017.

The FinnGen study is approved by Finnish Institute for Health and Welfare (permit numbers: THL/2031/6.02.00/2017, THL/1101/5.05.00/2017, THL/341/6.02.00/2018, THL/2222/6.02.00/2018, THL/283/6.02.00/2019, THL/1721/5.05.00/2019 and THL/1524/5.05.00/2020), Digital and population data service agency (permit numbers: VRK43431/2017-3, VRK/6909/2018-3, VRK/4415/2019-3), the Social Insurance Institution (permit numbers: KELA 58/522/2017, KELA 131/522/2018, KELA 70/522/2019, KELA 98/522/2019, KELA 134/522/2019, KELA 138/522/2019, KELA 2/522/2020, KELA 16/522/2020), Findata permit numbers THL/2364/14.02/2020, THL/4055/14.06.00/2020, THL/3433/14.06.00/2020, THL/4432/14.06/2020, THL/5189/14.06/2020, THL/5894/14.06.00/2020, THL/6619/14.06.00/2020, THL/209/14.06.00/2021, THL/688/14.06.00/2021, THL/1284/14.06.00/2021, THL/1965/14.06.00/2021, THL/5546/14.02.00/2020, THL/2658/14.06.00/2021, THL/4235/14.06.00/2021, Statistics Finland (permit numbers: TK-53-1041-17 and TK/143/07.03.00/2020 (earlier TK-53-90-20) TK/1735/07.03.00/2021, TK/3112/07.03.00/2021) and Finnish Registry for Kidney Diseases permission/extract from the meeting minutes on 4th July 2019.

The Biobank Access Decisions for FinnGen samples and data utilized in FinnGen Data Freeze 12 include: THL Biobank BB2017_55, BB2017_111, BB2018_19, BB_2018_34, BB_2018_67, BB2018_71, BB2019_7, BB2019_8, BB2019_26, BB2020_1, BB2021_65, Finnish Red Cross Blood Service Biobank 7.12.2017, Helsinki Biobank HUS/359/2017, HUS/248/2020, HUS/430/2021 §28, §29, HUS/150/2022 §12, §13, §14, §15, §16, §17, §18, §23, §58, §59, HUS/128/2023 §18, Auria Biobank AB17-5154 and amendment #1 (August 17 2020) and amendments BB_2021-0140, BB_2021-0156 (August 26 2021, Feb 2 2022), BB_2021-0169, BB_2021-0179, BB_2021-0161, AB20-5926 and amendment #1 (April 23 2020) and it’s modifications (Sep 22 2021), BB_2022-0262, BB_2022-0256, Biobank Borealis of Northern Finland_2017_1013, 2021_5010, 2021_5010 Amendment, 2021_5018, 2021_5018 Amendment, 2021_5015, 2021_5015 Amendment, 2021_5015 Amendment_2, 2021_5023, 2021_5023 Amendment, 2021_5023 Amendment_2, 2021_5017, 2021_5017 Amendment, 2022_6001, 2022_6001 Amendment, 2022_6006 Amendment, 2022_6006 Amendment, 2022_6006 Amendment_2, BB22-0067, 2022_0262, 2022_0262 Amendment, Biobank of Eastern Finland 1186/2018 and amendment 22§/2020, 53§/2021, 13§/2022, 14§/2022, 15§/2022, 27§/2022, 28§/2022, 29§/2022, 33§/2022, 35§/2022, 36§/2022, 37§/2022, 39§/2022, 7§/2023, 32§/2023, 33§/2023, 34§/2023, 35§/2023, 36§/2023, 37§/2023, 38§/2023, 39§/2023, 40§/2023, 41§/2023, Finnish Clinical Biobank Tampere MH0004 and amendments (21.02.2020 & 06.10.2020), BB2021-0140 8§/2021, 9§/2021, §9/2022, §10/2022, §12/2022, 13§/2022, §20/2022, §21/2022, §22/2022, §23/2022, 28§/2022, 29§/2022, 30§/2022, 31§/2022, 32§/2022, 38§/2022, 40§/2022, 42§/2022, 1§/2023, Central Finland Biobank 1-2017, BB_2021-0161, BB_2021-0169, BB_2021-0179, BB_2021-0170, BB_2022-0256, BB_2022-0262, BB22-0067, Decision allowing to continue data processing until 31st Aug 2024 for projects: BB_2021-0179, BB22-0067,BB_2022-0262, BB_2021-0170, BB_2021-0164, BB_2021-0161, and BB_2021-0169, and Terveystalo Biobank STB 2018001 and amendment 25th Aug 2020, Finnish Hematological Registry and Clinical Biobank decision 18th June 2021, Arctic biobank P0844: ARC_2021_1001.

#### Ethical Statement GEL

Ethical approval for the 100KGP was granted by the Research Ethics Committee for East of England – Cambridge South (REC Ref: 14/EE/1112). Participants provided written informed consent for the use of their genetic and clinical data.

### Clinical data collection

In IKGP Patients with a confirmed diagnosis of Autosomal Dominant Polycystic Kidney Disease (ADPKD) enrolled in the Irish Kidney Gene Project (IKGP) with available genotype data were included. Clinical information and imaging data were extracted from electronic healthcare records, including the E-Med system (the national renal information system), the Progeny pedigree database, and the National Integrated Medical Imaging System (NIMIS). Data collected included patient demographics, genetic results, comorbidities, renal status at last follow-up, family history of intracranial aneurysm (IA) or subarachnoid haemorrhage (SAH), and details of cerebral screening and any interventions performed.

Age, sex, and PKD genotype were obtained from the IKGP database and cross-checked against E-Med and clinic records. Family history of IA or SAH was ascertained from the medical record and verified against the family pedigree generated in Progeny at the time of IKGP enrolment; where a relative was subsequently diagnosed with an IA or SAH, the family history of all related participants was updated accordingly. Hypertension was defined as a documented diagnosis in E-Med, in outpatient clinic correspondence or in any other clinical document, and/or current treatment with one or more antihypertensive medications. Dyslipidaemia was defined using the same approach: a documented diagnosis in E-Med or clinical correspondence, and/or current treatment with lipid-lowering therapy. Diabetes mellitus was defined as a documented diagnosis in E-Med or clinical correspondence, and/or current treatment with glucose-lowering therapy (oral hypoglycaemic agents or insulin), non-diabetic patients receiving GLP-1 receptor agonists for weight loss, as well as those receiving SGLT2 inhibitors for other indications, were excluded. Smoking history was ascertained primarily from documentation in E-Med and clinic letters, and categorised as ever smoked, never smoked or unknown.

Renal status assessed at date of last follow-up,, defined as the earliest of last clinical contact, death, or the census date of 30 April 2025. It was determined from the estimated glomerular filtration rate (eGFR) recorded in E-Med, with chronic kidney disease stage assigned according to KDIGO criteria, alongside documentation of dialysis or transplant status. Cerebral screening was defined as any documented cerebral imaging performed to evaluate for intracranial aneurysm, including magnetic resonance angiography (MRA), computed tomography angiography (CTA), digital subtraction angiography (DSA), or conventional cerebral angiography; imaging studies and their formal radiology reports were reviewed via NIMIS. A diagnosis of IA was confirmed by review of the cerebral imaging, and only aneurysms measuring ≥2 mm were included. SAH events were identified from documented diagnoses in the patient record, with the corresponding imaging reviewed to confirm an aneurysmal source; non-aneurysmal SAH cases excluded.

In FinnGen release R13 (n=519,972, last date of follow up February 2025), polycystic kidney disease was ascertained with ICD-10 code Q61.2 from electronic health record data (causes of death register, hospital discharge register, primary care register, or drug reimbursement register). Harmonized smoking status information was available for 591/826 individuals through a combination of survey data from legacy cohorts included in the FinnGen cohort and smoking information provided by regional biobanks participating in the FinnGen project. For this analysis, we used the “ever smoker” variable representing whether an individual had ever engaged in smoking in their lifetime. Sex was genetically determined from genotyping data. Strict criteria were used to define IA or SAH in FinnGen (details below).

Dyslipidaemia was ascertained with FinnGen endpoint E4_HYPERCHOL (ICD-10 E78.0, ICD-9 2720, ICD-8 2720 from hospital discharge or cause of death registry). Hypertension was ascertained with FinnGen endpoint I9_HYPTENS (ICD-10 I10-I15, I67.4, ICD-9 4019X|4029A|4029B|4039A|4040A|4059A|4059B|4372A|4059X, ICD-8 400|401|402|403|404 from hospital discharge or cause of death registry, or KELA code 205 from KELA reimbursement registry). Diabetes was ascertained with endpoint E4_DIABETES (ICD-10 E10 E11). For each phenotype, age and date of diagnosis was taken as first appearance of the code in that individual.

Family history was ascertained by finding individuals within the FinnGen cohort that are genetically related to the case, which was previously calculated by the FinnGen core team with King relatedness inference^62^. If an ADPKD case had a genetically related individual (up to 3^rd^ degree) with an I9_ANEURYSM, I9_ANEUR, or I9_SAH diagnosis, they were considered having a positive family history of SAH or IA. End-stage renal failure was ascertained from first diagnosis of FinnGen core endpoint DIALYSIS (ICD-10 E11 from hospital discharge or cause of death registry, ATC code A10B from medicine purchases registry, minimum 3 events), Z21_CARE_INVOLVI_DIALY (ICD-10 Z49 from hospital discharge or cause of death registry), Z21_EXTRACO_DIALY (ICD-10 Z49.1 from hospital discharge or cause of death registry), Z21_OTHER_DIALY (ICD-10 Z49.2 from hospital discharge or cause of death registry), N14_DIALYSIS (ICD-10 Y84.1 or Z99.2 from hospital discharge or cause of death registry, KELA code 137 from KELA reimbursements registry), or Z21_KIDNEY_TRANSLPLANT_STATUS (ICD-10 Z94.0 from hospital discharge or cause of death registry).

In GEL, cystic kidney disease patients were included if they had more than five renal cysts affecting one or both kidneys in combination with atypical features (including non-classical imaging appearances, onset before age 10, syndromic features, or where a genetic diagnosis would influence management), or features consistent with classical ADPKD without prior genetic testing of *PKD1* or *PKD2*; those with non-cystic causes of kidney failure, multicystic dysplastic kidneys, or prior genetic diagnoses were excluded. Linked Hospital Episode Statistics records were interrogated to derive clinical outcomes and comorbidities, including kidney failure and chronic kidney disease stage, with age at kidney failure defined by the earliest relevant diagnostic code; the same approach was applied to identify intracranial aneurysm and subarachnoid haemorrhage events. Additional clinical variables, including diabetes, dyslipidaemia, hypertension, smoking status, and family history of subarachnoid haemorrhage, were extracted using predefined coding frameworks (Supplementary Data 3). Demographic characteristics were obtained from LabKey tables associated with participant records. Data from GEL were from Release v19 (31/10/2024).

### Additional criteria for SAH definition in FinnGen

SAH cases were identified using the Finnish the Care Register for Health Care, which covers inpatient and specialist outpatient contacts, and the Causes of Death Register. Contacts recorded only in the primary health care register (Avohilmo) were not used, because diagnostic coding there was judged too unreliable for identifying an acute event like SAH.

We began with every FinnGen participant who had ICD-10 code I60.0–I60.6 recorded as a primary or secondary diagnosis in a hospital contact, or as the underlying cause of death, from 1998 onward. Anyone who also had diagnosis of traumatic brain injury (S06.0–S06.9) or a cerebral arteriovenous malformation (Q28.0–Q28.9) was removed from this candidate group, since these suggest the SAH was not a spontaneous aneurysmal event.

Some candidates died of SAH without ever having a hospital contact coded for it. These sudden, out-of-hospital deaths were included in the case set directly, without being checked against the rules described below.

Because FinnGen’s data do not include a direct indicator for whether an admission was an emergency, we defined emergency admission using two signals: either the admission itself was coded as urgent, or the patient had an emergency-department visit that led to hospital admission on the same day. This definition applies everywhere emergency admission is mentioned below. Where possible, we also cross-checked that a patient’s neurosurgical admission was genuine by looking for a matching neurosurgical procedure code recorded from three days before to thirty days after their first hospital admission.

Candidates with a qualifying index event, an emergency admission with SAH as the primary diagnosis and no nearby trauma or AVM code were then checked against a series of rules in order and classified according to the first rule they matched. For candidates whose neurosurgical admission could be confirmed by a matching operation (NOMESCO codes AAC00, AAC10, AAC20, AAL00, PA2KT, PA2LT, or PA2MT), we first checked whether the admission itself was an emergency admission straight to neurosurgery; failing that, whether an emergency-department visit was followed by neurosurgical admission on the same day; failing that, whether an emergency-department visit was followed by neurosurgical admission one to three days later; failing that, whether the patient had a neurosurgical admission with no positive urgency signal but also no explicit non-urgent code; and finally, whether any other admission lacking both an urgency signal and a neurosurgery code was nonetheless supported by the matched operation alone.

For candidates whose neurosurgical admission could not be confirmed by a matching operation, we checked a parallel series of rules in the same way. We first checked for an emergency admission straight to neurosurgery, then for an emergency-department visit followed by same-day neurosurgical admission, then for a transfer from a non-neurosurgical to a neurosurgical admission within thirty days of the index event, then for death from SAH within one year of the index event with a positive urgency signal, and finally for death from SAH within one year of the index event without a positive urgency signal.

Candidates who matched none of these rules were excluded if they died of a non-SAH cause within one year of the index event, or if their index hospitalization lasted fewer than ten days. A hospitalization of ten days or more, on its own, was judged too weak a signal to justify inclusion.

Candidates who never had a qualifying index event were checked against one further rule: emergency admission to neurosurgery with unspecified SAH (I60.7– I60.9) as the primary diagnosis and aneurysmal SAH (I60.0–I60.6) as a secondary diagnosis, with no nearby trauma or AVM code.

### Additional criteria for intracranial aneurysm diagnosis in FinnGen

To ensure accurate phenotyping from the EHR-based FinnGen data, we applied further filtering to intracranial aneurysm diagnoses in FinnGen. An intracranial aneurysm diagnosis was defined as the first recorded ICD-10 code I67.1 from 1998 onwards, as ICD-10 codes were used inconsistently in the Finnish healthcare system prior to this year. The date of this first I67.1 entry was used as the index date (Day 0) for each patient. To ensure that the diagnosis reflected a true unruptured intracranial aneurysm rather than a ruptured lesion or a non-aneurysmal cerebrovascular condition, two exclusion criteria were applied. Patients with any diagnosis of subarachnoid or other ruptured intracranial haemorrhage (ICD-10 codes I60.0–I60.9) recorded at any point in their available medical history up to and including 90 days after the index date were excluded. Patients with a diagnosis of congenital cerebrovascular malformation or other specified cerebrovascular disease (ICD-10 codes Q28.0–Q28.9 or I67.5) recorded at any point in their available history up to and including 90 days after the index date were also excluded. Next, each remaining patient was required to have documentation of either an aneurysm-directed treatment or a relevant diagnostic imaging procedure occurring between 365 days before and 90 days after the index date. Patients with no qualifying treatment or imaging code within this window were not considered to have a confirmed aneurysm diagnosis.

### Classification of treatment status

In FinnGen, patients were classified as having received aneurysm treatment if any of the following NOMESCO Classification of Surgical Procedures (NCSP) codes were recorded within −365 to +90 days of the index date: AAC00, AAC10, AAC20, AAL00, PA2KT, PA2LT, PA2MT, or PA6YT. The PA6YT code was applied only to patients with no previous diagnosis of I65.2 or I63.2. Patients meeting any of these criteria were assigned to the aneurysm treatment group.

### Classification of imaging modality

Patients not assigned to the treatment group were further classified according to the most specific diagnostic imaging procedure recorded within −365 to +90 days of the index date, using a hierarchical algorithm.

Patients were first assessed for cerebral digital subtraction angiography (DSA), defined by procedure codes PA2AC, PA2BC, PA2CC, or PA7CC. Those meeting any of these criteria were assigned to the cerebral DSA group. The remaining patients were assessed for head and neck CT angiography (CTA) or magnetic resonance angiography (MRA), defined by codes PA2AD, PA2BD, PA2AM, PA2BM, PA2CM, PA2AG, PA2BG, PA2CG, PA2DG, PA7AD, PA7BD, or PA7CD, and assigned to the head and neck CTA/MRA group. The remaining patients were assessed for neck CTA (codes PA3AC, PA4AC, PA6AD, or PA6BD) and assigned to the neck CTA group. The remaining patients were assessed for plain brain MRI (codes AA1AG, AA1AM, AA1BG, AA1BM, AA1CG, AA1CM, AA1DG, or AA1DM) and assigned to the plain brain MRI group. Finally, any remaining patients were assessed for neck MRA only (codes PA6AG, PA6AM, PA6BG, or PA6CG) and assigned to the neck MRA only group. At each step, patients meeting the criteria were removed from further classification, ensuring mutually exclusive group assignment.

### Smoking status imputation

#### Smoking imputation in FinnGen

In FinnGen, smoking status (missing in 22.5%) was imputed by multiple imputation by chained equations (MICE)^63^ in R. To maximise the information available for the imputation model, smoking status was imputed across the entire FinnGen cohort (n= 519972) prior to sub-setting to ADPKD patients. Smoking was treated as a binary variable and imputed using logistic regression, with the imputation model including age, sex, hypertension and dyslipidaemia as predictors. All other variables were explicitly excluded from the imputation model. A total number of 25 imputed datasets were generated over 25 iterations. To enable a unified analysis across imputations, the twenty-five imputed datasets were combined into a single stacked dataset, where imputed smoking status varied but all other phenotypes were constant between individual patient replicates. Each row was assigned a frequency weight of 1/m (where m = 25), preserving the effective sample size of the original cohort. The post-imputation smoking prevalence in ADPKD patients was ∼49.2%, closely matching the pre-imputation observed prevalence of ∼49.2% in those with no missing values, supporting the plausibility of the imputed values. All subsequent multivariate analyses were performed on this stacked dataset using cluster-robust standard errors with the original patient identifier as the clustering variable, accounting for the non-independence of the m replicate rows per patient. Stacking with frequency weights and cluster-robust variance was used in preference to Rubin’s-rules pooling because the standard implementation of the pooled likelihood ratio test for nested Cox models was not numerically stable with our model specification.

#### Smoking imputation in IKGP

In the genotyped IKGP cohort of 475 patients, smoking status had originally been missing in 291/475 patients (61.3%), with an observed pre-imputation smoking prevalence of 27.7% (51/184) among those with available data.

Smoking imputation was performed across the 356 IKGP ADPKD patients with known cerebral screening status using multiple imputation by chained equations (MICE) in STATA 19 via the mi impute chained command^64^. Smoking was treated as a binary variable and imputed using logistic regression, with the imputation model including age, sex, hypertension, and dyslipidaemia as predictors. All other variables, including the outcome (intracranial aneurysm status), were explicitly excluded from the imputation model to avoid circularity. A total of 20 imputed datasets were generated.

Of the 475 genotyped patients eligible for the current study, 326 met the criteria for inclusion in the smoking imputation set of whom 141 had originally observed smoking data and 185 underwent imputation. The remaining genotyped patients without documented screening status did not undergo imputation. The post-imputation smoking prevalence in the analysed cohort was 27.5%, closely matching the pre-imputation observed prevalence and supporting the plausibility of the imputed values. The imputed smoking status was incorporated as a covariate in the logistic regression model, which was fitted separately on each imputed dataset using mi estimate. Results were pooled across imputed datasets using Rubin’s rules^65^.

Formal sensitivity analyses, including bootstrap stability testing and cross-validation, could not be performed due to the limited sample size; nonetheless, the close concordance between the observed and imputed smoking distributions, the use of clinically relevant predictors, and the consistent direction of effect across alternative imputation approaches support the plausibility of the imputed values.

#### Smoking imputation in GEL

In GEL, smoking status was ascertained via ICD codes in electronic health record data. Imputation was not undertaken in this cohort, and analyses involving smoking were performed as complete-case, and the resulting estimates should be interpreted with this in mind.

### Monogenic variant evaluation

Monogenic variants in IKGP were evaluated by using a combination of next-generation sequencing strategies, including whole-exome sequencing and targeted kidney gene panels covering known cystic and chronic kidney disease genes. Long-range PCR was used to address the pseudogene-rich region of *PKD1*, and *MUC1* sequencing with immunostaining was performed where clinically indicated. Variants were classified according to the American College of Medical Genetics and Genomics (ACMG) guidelines, with the Mayo Clinic pathogenicity criteria additionally applied to *PKD1* and *PKD2* variants. Only variants classified as pathogenic or likely pathogenic were considered disease-causing^66^.

Monogenic variants in FinnGen were evaluated with whole-exome sequencing, performed at the Institute for Molecular Medicine Finland (FIMM) Genomics Unit. 50 ng of gDNA was processed according to Twist Library Preparation EF 2.0 with Enzymatic Fragmentation DOC-001239 REV 1.0 and Twist Target Enrichment Protocol DOC-001273 REV 3.0 manual (Twist Bioscience, San Francisco, CA, USA) with following modifications. 7 PCR cycles were used in library preparation. Following adapters were used for ligation: Twist Full Length UDI adapters (Twist Bioscience, San Francisco, CA, USA). Library quantification and quality check was performed using 5300 Fragment Analyzer system HS NGS Fragment Kit (Agilent, Santa Clara, CA, USA) and Qubit Broad Range DNA Assay (Thermo Fisher Scientific, Waltham, MA, USA). Libraries were pooled to 8 or 7 –plex reactions according to concentration (Qubit BR assay). The exome enrichment was performed using Twist Exome 2.0 + Comp.Exome spike-in probes (37,45 Mb). The captured library pools were quantified for sequencing using QuantStudio 5 Collibri Library Quantification kit (Thermo Fisher Scientific, Waltham, MA, USA) and either Agilent Bioanalyzer 2100 High Sensitivity assay (Agilent, Santa Clara, CA, USA) or 5300 Fragment Analyzer system HS NGS Fragment Kit (Agilent, Santa Clara, CA, USA). Sequencing was performed with Illumina NovaSeq 6000 system using S4 flow cell (Illumina, San Diego, CA, USA) and v1.5 chemistry. Read length for the paired-end run was 101+10+10+101bp.

Variant calling was performed with Illumina DRAGEN v4.2.9, followed by further quality control of exome sequencing data. Briefly, samples with contamination greater than 5% were excluded (1 sample), and samples were checked to ensure that all samples fell within 4 standard deviations of the batch mean on transition/transversion ratio, insertion/deletion ratio, and number of singletons. Genotypes were filtered out if their read depth was less than 10 or Phred likelihood (PL) of the reference call was less than 20, or if their fraction of reference reads was less than 90% (for homozygous variant genotype calls) or outside 20-80% (for heterozygous genotype calls). Variants failing DRAGEN hard filters were removed, as well as those where >20% of heterozygous genotype calls had a fraction of reference reads outside 20-80%.

Passing variants were annotated with Variant Effect Predictor (VEP)^67^ v.95 with the LOFTEE^68^ plugin, gnomAD^68^ v2 allele frequencies (including whether the variant had significantly different allele frequencies in gnomAD genomes vs exomes datasets), CADD^69^ v1.6 Phred scores, and ClinVar^56^ release from February 26^th^, 2026. Variants were considered likely related to disease if they were LOFTEE high-confidence predicted loss-of-function variants or missense variants with CADD Phred score >20 and rare in global populations in the genes *PKD1*, *PKD2*, *PKHD1*, *GANAB*, *IFT140*, *ALG8, ALG5, ALG9,* or *DNAJB11*. Variants were considered rare in global populations if they were missing from gnomAD, or present with allele count <10 in all populations and no reported homozygous carriers (for heterozygous variants) or present with allele frequency <1% in all populations and fewer than 10 homozygous carriers in all populations. In addition to in-silico predicted likely damaging variants, we considered all variants annotated in ClinVar as pathogenic or likely pathogenic with disease matching all or in part ‘Polycystic kidney disease’ or in the genes *PKD1, PKD2, PKHD1, GANAB, IFT140, ALG8, ALG5, ALG9,* or *DNAJB11*. Variants were considered truncating if they were high-confidence LOFTEE predicted loss-of-function variants or annotated as splice acceptor, splice donor, nonsense, or frameshift variants by ClinVar, and considered non-truncating if variants were annotated as VEP or ClinVar as missense or inframe deletion variants.

Monogenic variants in GEL were prioritised for pathogenicity using distinct criteria for truncating and non-truncating classes within a gene-based collapsing framework. Truncating variants were defined as high-confidence loss-of-function events, including stop-gained, frameshift, and essential splice site variants, as annotated by the LOFTEE^68^ plugin in the Ensembl Variant Effect Predictor^67^ with high confidence, and were restricted to rare variants (MAF <0.01% in gnomAD^68^). Non-truncating variants comprised rare coding variants predicted to be damaging, including missense, in-frame insertions/deletions, and start-loss variants, selected using a deleteriousness threshold of CADD^69^ ≥20 to enrich for the top 1% of predicted functional impact. Across both classes, variants were required to meet stringent quality control thresholds, including low allele count (MAC ≤20), adequate sequencing depth, high genotype quality, and minimal missingness, to ensure robustness of downstream association analyses, as previously reported^61^.

### Genotyping and Quality Control

In IKGP, samples were genotyped on the Infinium™ OmniExpressExome-8 chip (Illumina Inc., San Diego, CA, USA). PLINK (ver. 1.9) was used to perform quality control on the data. Only autosomal variants were included for downstream analysis. Variants with a Hardy–Weinberg equilibrium p-value < 1×10⁻, with missingness >5%, and a minor allele frequency <2% were excluded. Samples with >5% SNP missingness were also excluded. Post-QC, the data was submitted to the Sanger server for imputation with the Haplotype Reference Consortium (r1.1) panel and was phased with EAGLE2 and PBWT tools pre-imputation.

Genotyping and quality control in FinnGen were performed centrally by the FinnGen core analysis team. Samples were genotyped using Illumina (Illumina Inc., San Diego, CA, USA) and Affymetrix (Thermo Fisher Scientific, Santa Clara, CA, USA) arrays. Genotype calling was conducted using the GenCall and zCall algorithms for Illumina data, and the AxiomGT1 algorithm for Affymetrix data.

Standard quality control procedures were applied to exclude individuals with ambiguous sex, genotype missingness >5%, excess heterozygosity (±4 standard deviations from the mean), or non-Finnish ancestry. Variants were excluded if they had >2% missingness, a Hardy–Weinberg equilibrium p-value < 1×10⁻, or a minor allele count (MAC) < 3. Pre-phasing of array data was performed with Eagle v2.3.5, using 20,000 conditioning haplotypes. Further details on the genotyping and imputation pipelines are available in the FinnGen flagship paper^54^.

Whole genome sequencing and primary processing in the 100,000 Genomes Project were performed centrally by Genomics England. Germline DNA was sequenced using 150 base pair paired end reads on Illumina HiSeq X instruments and processed using the Illumina North Star Version 4 workflow v2.6.53.23, with alignment to the GRCh38 reference genome including decoy sequences using iSAAC v03.16.02.19 and small variant calling using Starling v2.4.7. Common variants were extracted from the Genomics England AggV2 data set, comprising jointly genotyped gVCFs from 78,195 quality controlled germline genomes. Joint genotyping was performed using Illumina gVCF Genotyper v2019.02.26, followed by variant normalisation, left alignment, and decomposition into biallelic representations using vt v0.57721. Following participant quality control, relatedness filtering, and ancestry matching, autosomal single nucleotide variants and indels were retained if they had a minor allele frequency of at least 0.1%, minor allele count of at least 20, missingness below 1%, Hardy Weinberg equilibrium P greater than 1 × 10, and differential missingness P greater than 1 × 10.

### Polygenic Risk Score Calculation

For polygenic risk scores based on unconditioned GWAS summary statistics, we followed typical PRS calculation methods. First, SNP weights for PRS construction were estimated using PRS-CS^70^, using the UK Biobank European LD reference panel (IKGP, GEL) or the Finnish-specific SiSu LD panel (FinnGen). Polygenic risk scores were then calculated with plink 1.9^71^ –-score sum, followed by z-scores standardization with the R^72^ v.4.5.3 scale () function. Polygenic risk scores were then calculated with plink 1.9^71^ –-score sum, followed by z-scores standardization with the R^72^ scale () function.

For polygenic risk scores based on conditioned GWAS summary statistics, we first conditioned the effect sizes of each phenotype’s genetic variants on the remaining two phenotypes with mtCOJO^58^. In other words, the conditioned effect sizes for SNPs associated with intracranial aneurysm from Bakker et al^27^ reflect the effect on IA after accounting for their effect on smoking intensity (cigarettes per day^29^) and on hypertension^59^. After conditioning each phenotype’s GWAS summary statistic on the remaining two phenotypes, we followed an identical protocol as above– SNP weight estimation with PRS-CS, scoring each individual, and z-score standardization.

### Cumulative incidence curves meta-analysis

A pooled standard error was computed across the three cohorts by first recovering stratum-specific variances from the 95% confidence intervals of the Kaplan-Meier estimates (using the normal approximation: SE = CI width / 3.92). These variances were then combined as a risk-set-weighted average, weighting each cohort’s variance by its risk set size at each time point, and the square root was taken to yield the pooled standard error. P value for each survival curve stratified by polygenic score represents the significance of the top 20% of polygenic risk compared to bottom 20% of polygenic risk in a Cox proportional hazards model (coxph, R survival package) with time-to-event defined via Surv(time, IA or SAH), with no further covariates in the model. P-values in the pooled survival curves across all three cohorts represent a random-effects meta-analysis (metagen, R meta package) of the effect of the top 20% of polygenic risk compared to the bottom 20% of polygenic risk in each cohort.

Survival curves were also estimated with the same method for monogenic variant carrier status, stratifying on *PKD1* truncating or non-truncating variant carriers, *PKD2* truncating or non-truncating variant carriers, or no variant found (removing individuals from the analysis carrying rare variants other than in *PKD1* or *PKD2*). Survival curves were estimated independently in each cohort, and pooled survival curves were calculated with the method described in above.

## Supplementary Figures

**Supplementary Figure 1.**
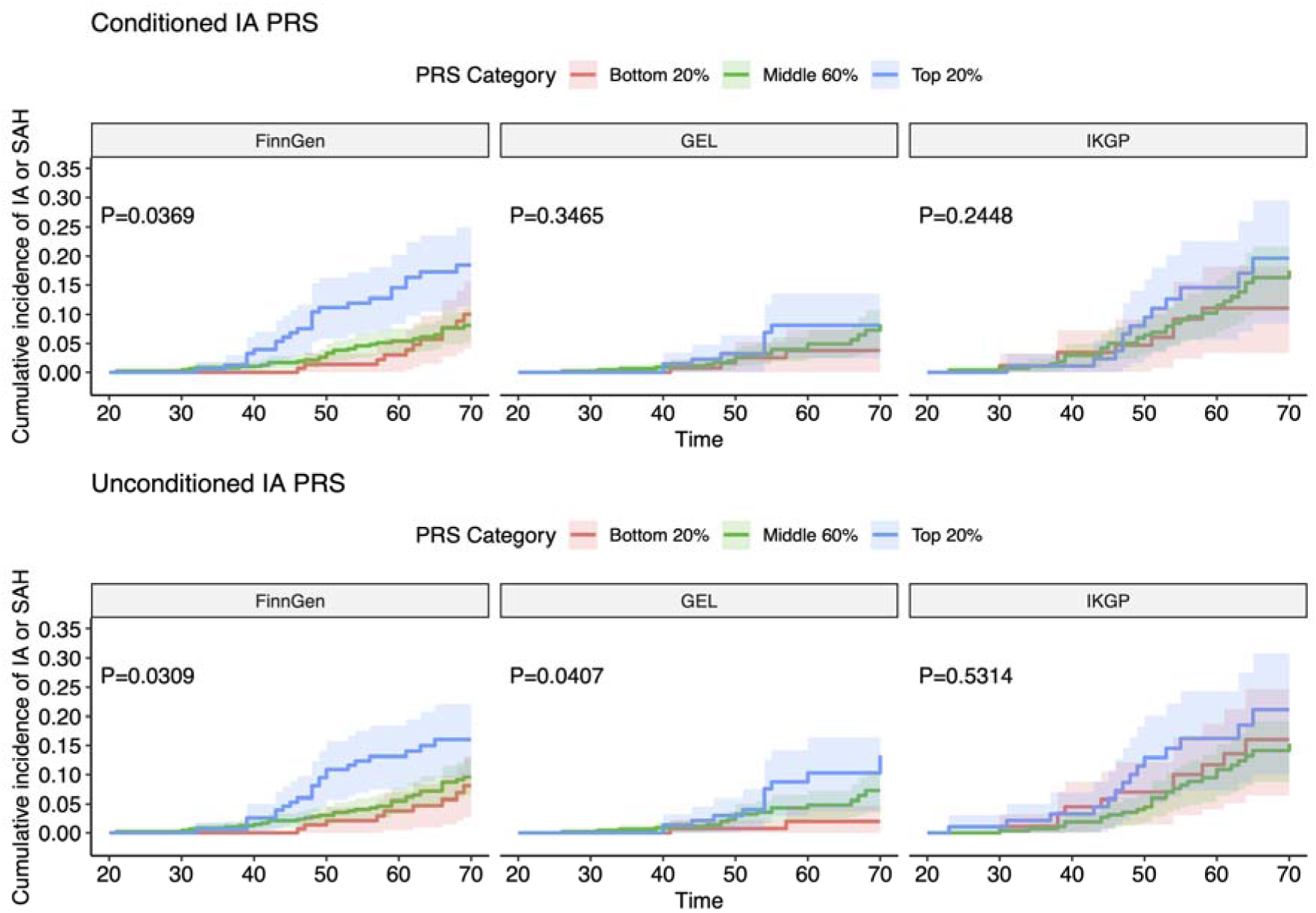
Intracranial aneurysm polygenic risk stratifying IA or SAH in each cohort. Cumulative incidence of IA or SAH by intracranial aneurysm (IA) PRS, with the top row representing polygenic scores calculated on mtCOJO conditioned GWAS summary statistics and the bottom row representing polygenic scores calculated from original unconditioned GWAS summary statistics. P-value represents categorical Cox univariate regression with 10 first principal components of genetic information as covariates, comparing top 20% of polygenic score risk to bottom 20% of polygenic score risk in each cohort and PRS type (conditioned, unconditioned).

**Supplementary Figure 2.**
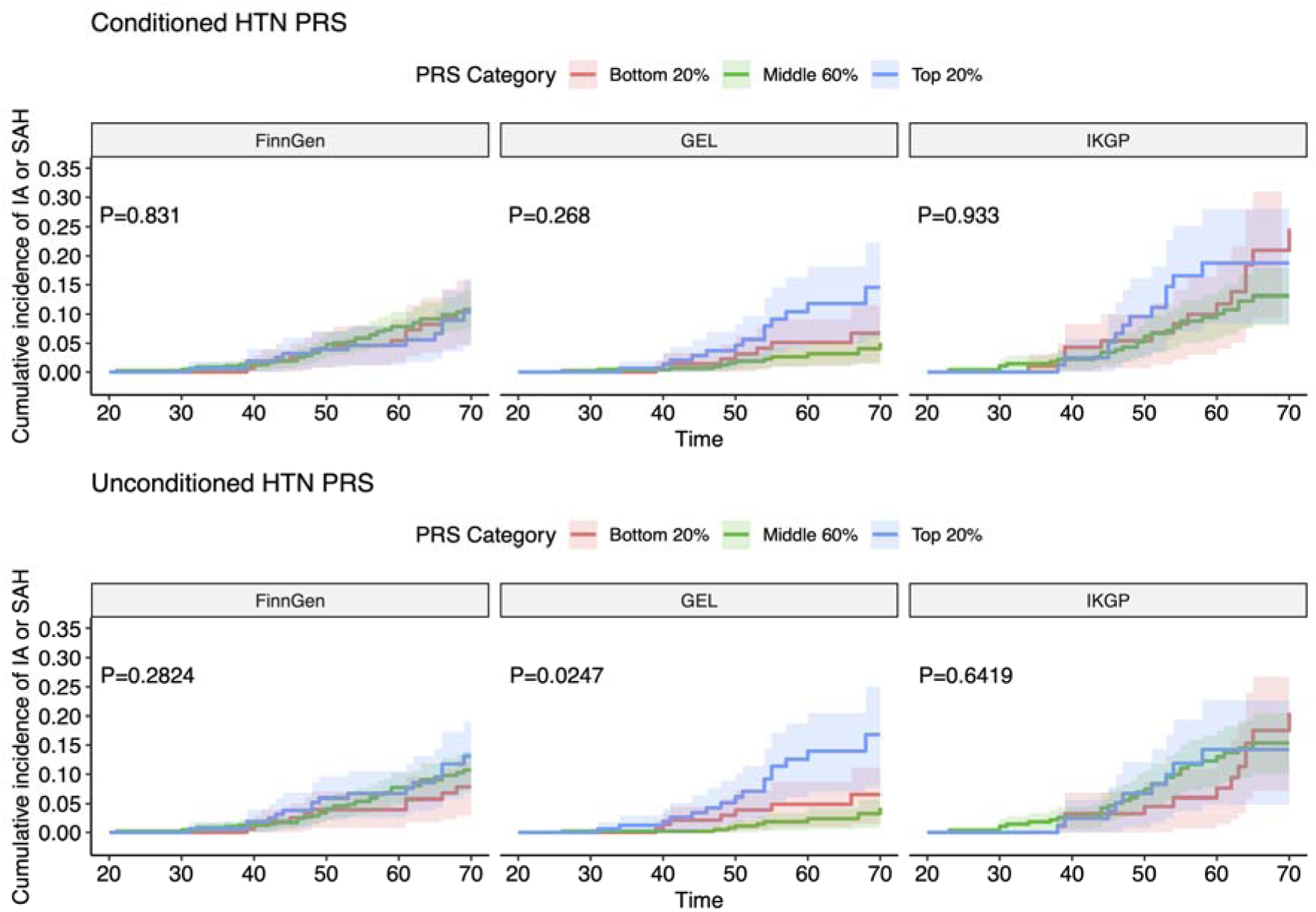
Hypertension polygenic risk stratifying IA or SAH in each cohort. Cumulative incidence of IA or SAH by hypertension PRS, with the top row representing polygenic scores calculated on mtCOJO conditioned GWAS summary statistics and the bottom row representing polygenic scores calculated from original unconditioned GWAS summary statistics. P-value represents categorical Cox univariate regression with 10 first principal components of genetic information as covariates, comparing top 20% of polygenic score risk to bottom 20% of polygenic score risk in each cohort and PRS type (conditioned, unconditioned).

**Supplementary Figure 3.**
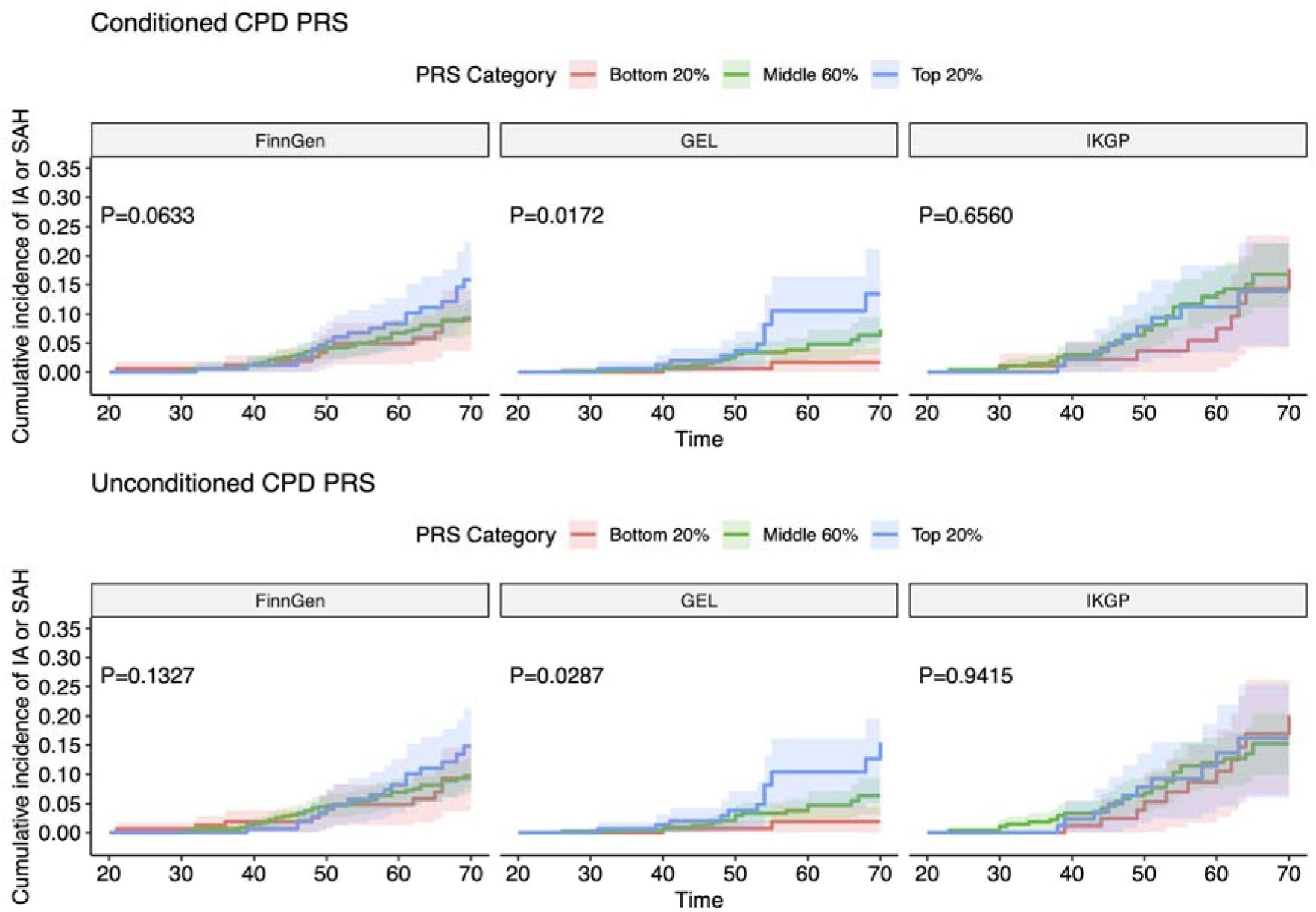
Smoking intensity polygenic risk stratifying IA or SAH in each cohort. Cumulative incidence of IA or SAH by smoking intensity (cigarettes per day, CPD) PRS, with the top row representing polygenic scores calculated on mtCOJO conditioned GWAS summary statistics and the bottom row representing polygenic scores calculated from original unconditioned GWAS summary statistics. P-value represents categorical Cox univariate regression with 10 first principal components of genetic information as covariates, comparing top 20% of polygenic score risk to bottom 20% of polygenic score risk in each cohort and PRS type (conditioned, unconditioned).

**Supplementary Figure 4.**
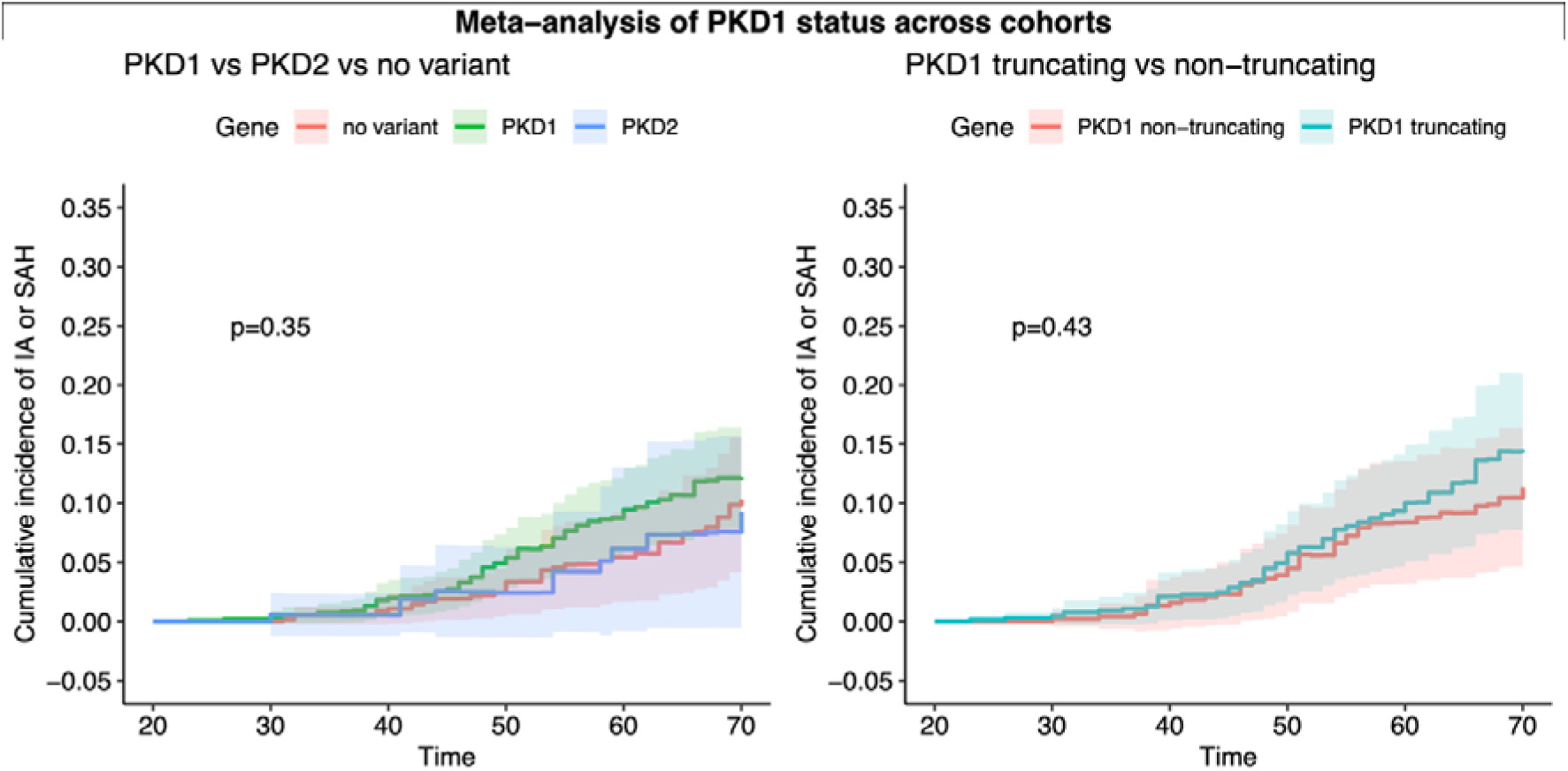
Cumulative incidence of IA or SAH by monogenic variant status, meta-analysis. Cumulative incidence of IA or SAH by *PKD1*, *PKD2*, or no variant found (left) and *PKD1* truncating vs non-truncating (right). P-value represents inverse-variance weighted meta-analysis of categorical Cox univariate regression with 10 first PCs as covariates across cohorts, comparing PKD1 vs no variant (left) and PKD1 truncating vs non-truncating (right).

**Supplementary Figure 5.**
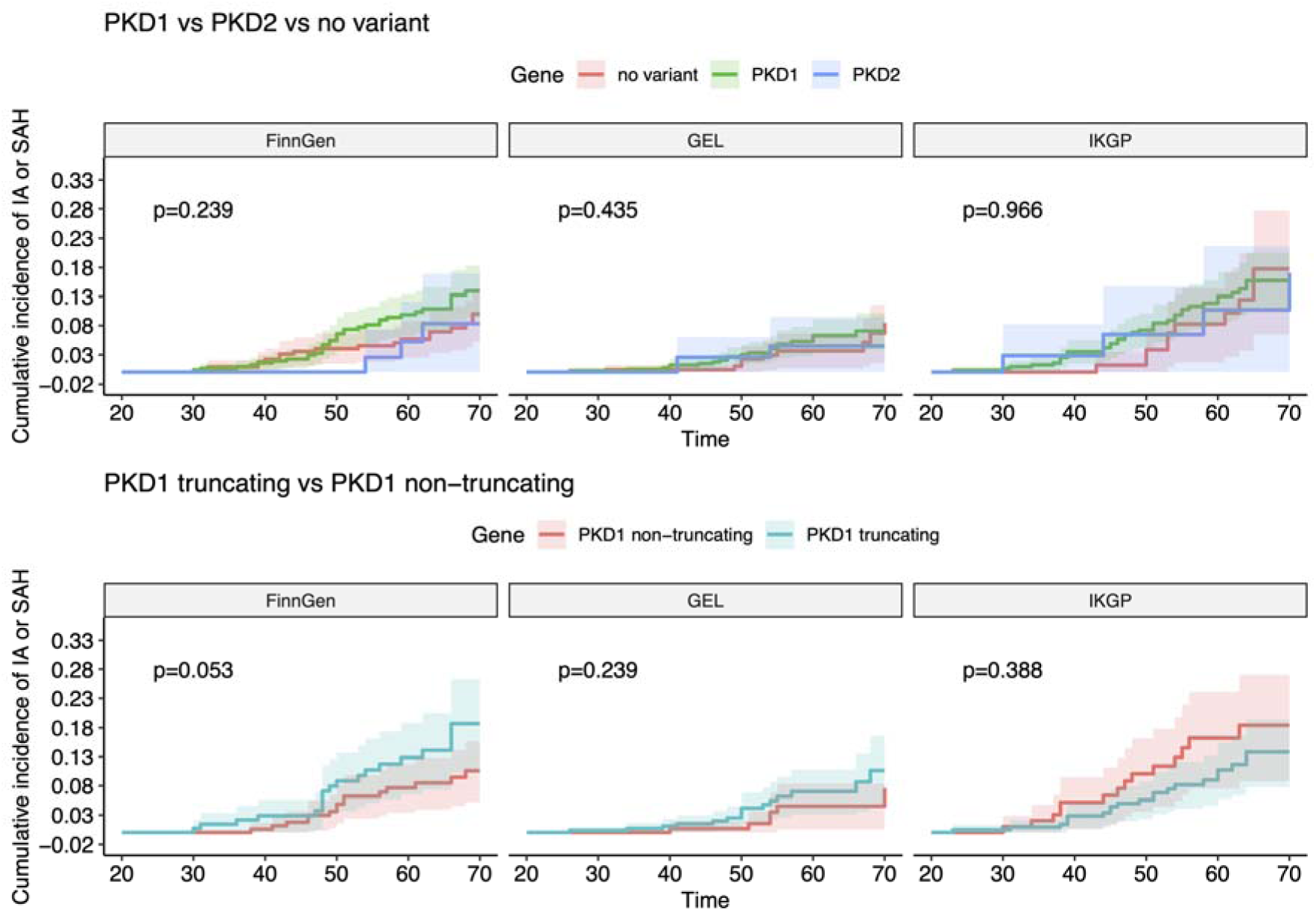
Cumulative incidence of IA or SAH by monogenic variant status, per cohort. Cumulative incidence of IA or SAH by *PKD1*, *PKD2*, or no variant found (top row) and *PKD1* truncating vs non-truncating (bottom row). P-value represents categorical Cox univariate regression with 10 first PCs as covariates across cohorts, comparing PKD1 vs no variant (top row) and PKD1 truncating vs non-truncating (bottom row).

**Supplementary Figure 6.**
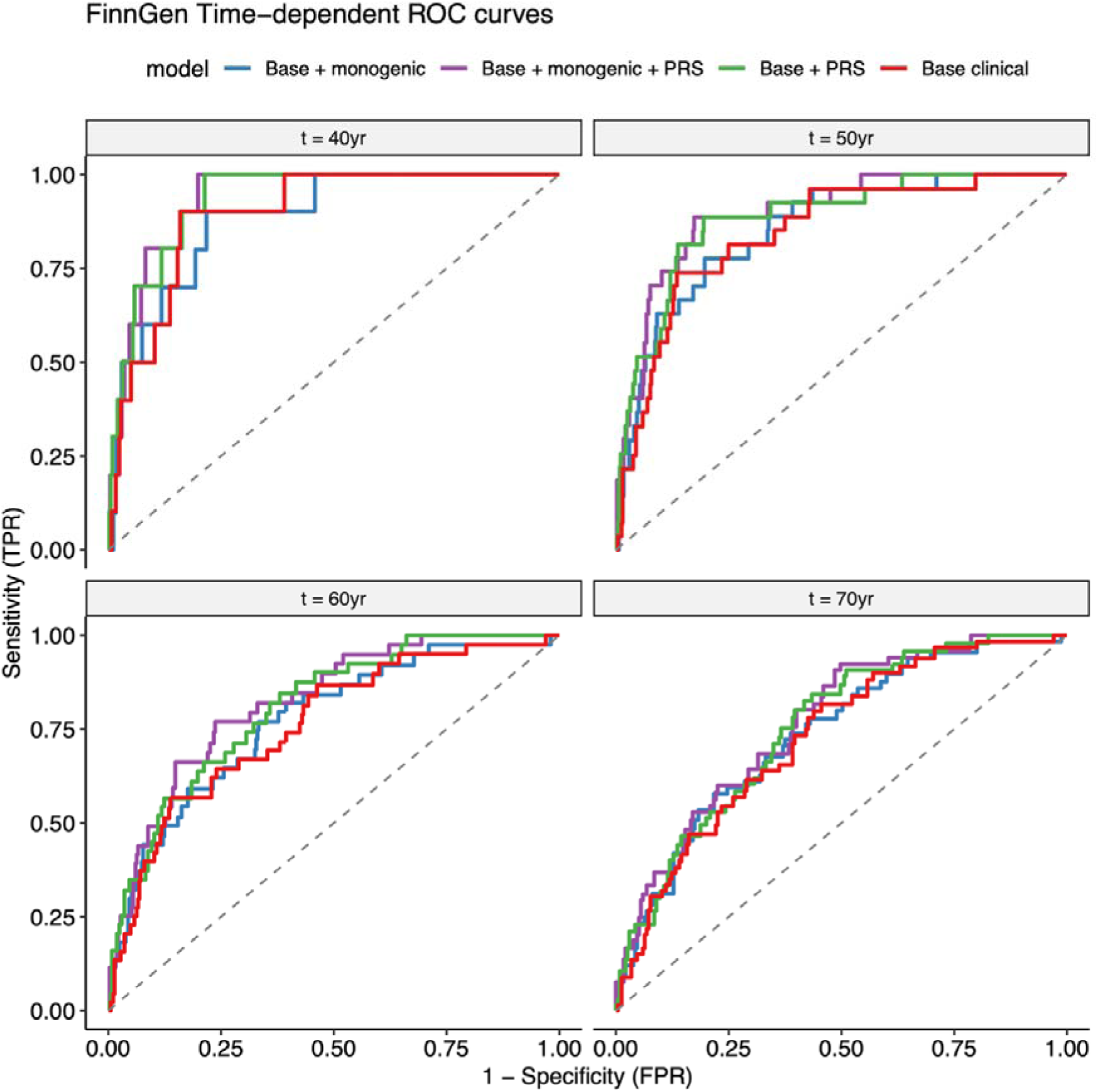
FinnGen time-dependent ROC curves. Time-dependent ROC curves for four models (Base clinical, Base + PRS, Base + Monogenic, Base + PRS + Monogenic) predicting intracranial aneurysm or subarachnoid haemorrhage, estimated using the timeROC package in R at 40, 50, 60, and 70 years of age. Each curve plots sensitivity against 1 – specificity.

**Supplementary Figure 7.**
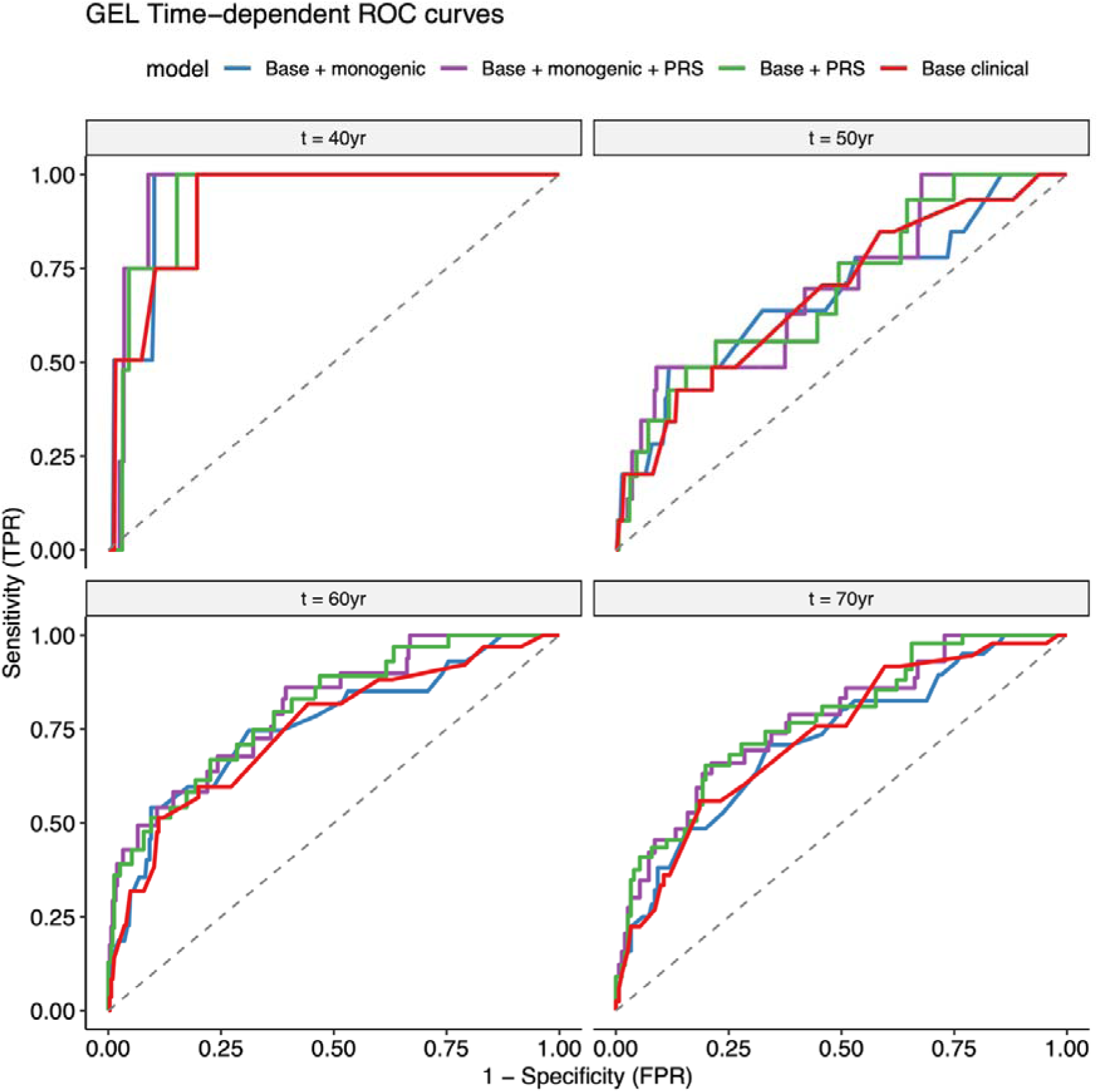
GEL time-dependent ROC curves. Time-dependent ROC curves for four models (Base clinical, Base + PRS, Base + Monogenic, Base + PRS + Monogenic) predicting intracranial aneurysm or subarachnoid haemorrhage, estimated using the timeROC package in R at 40, 50, 60, and 70 years of age. Each curve plots sensitivity against 1 – specificity.

**Supplementary Figure 8.**
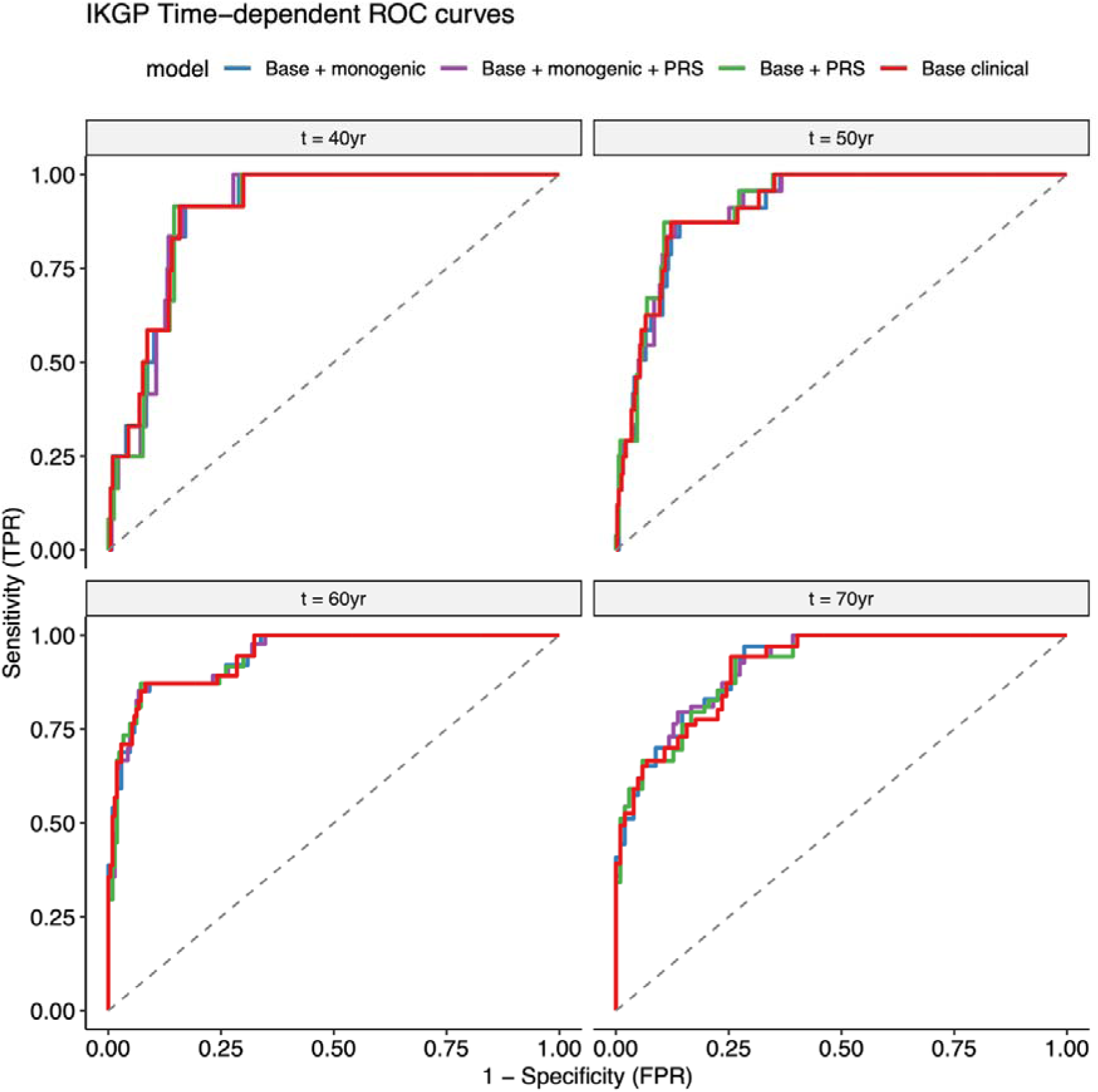
IKGP time-dependent ROC curves. Time-dependent ROC curves for four models (Base clinical, Base + PRS, Base + Monogenic, Base + PRS + Monogenic) predicting intracranial aneurysm or subarachnoid haemorrhage, estimated using the timeROC package in R at 40, 50, 60, and 70 years of age. Each curve plots sensitivity against 1 – specificity.

**Supplementary Figure 9.**
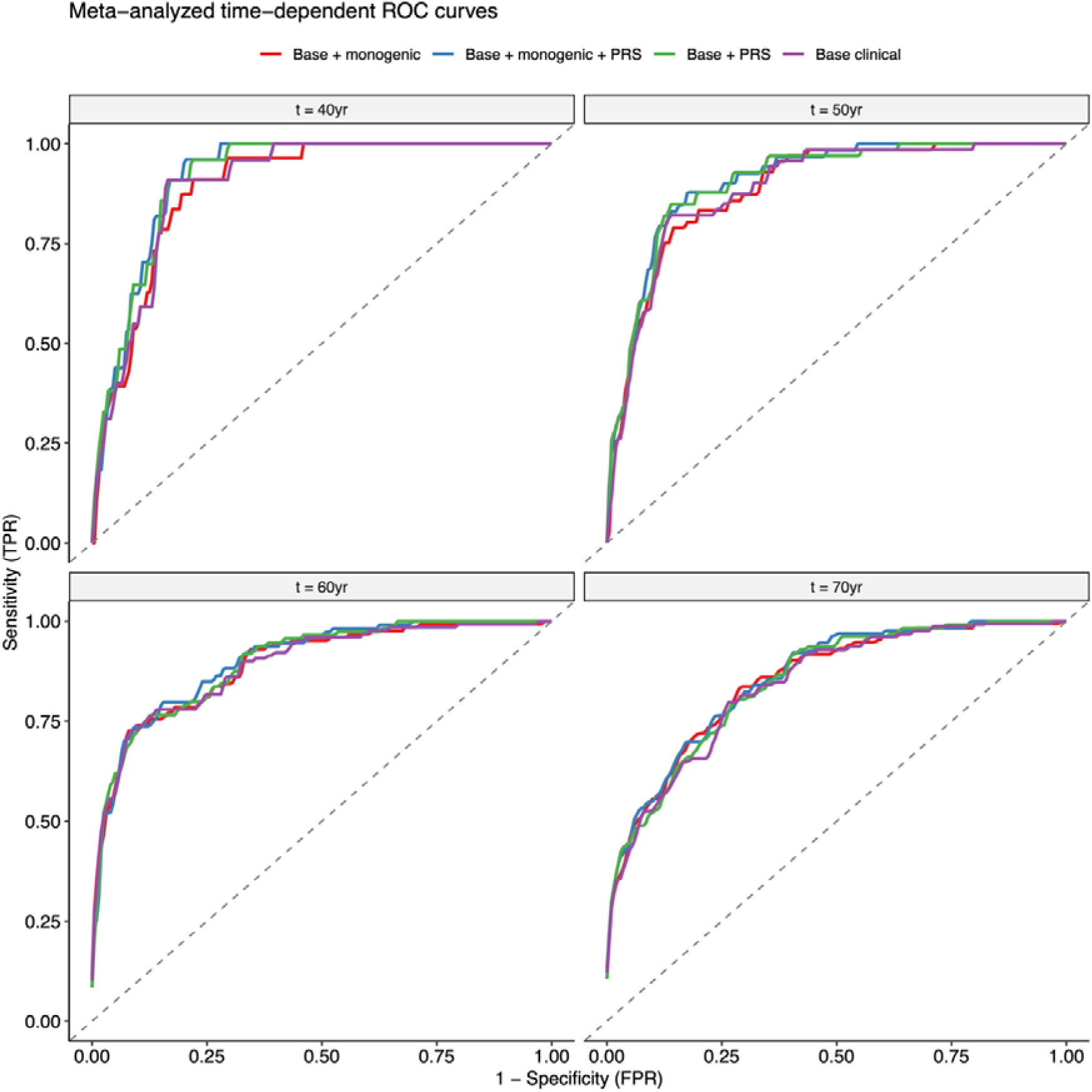
Meta-analyzed time-dependent ROC curves. Inverse-variance weighted meta-analysis of time-dependent ROC curves for four models (Base clinical, Base + PRS, Base + Monogenic, Base + PRS + Monogenic) predicting intracranial aneurysm or subarachnoid haemorrhage, estimated using the timeROC package in R at 40, 50, 60, and 70 years of age. Each curve plots sensitivity against 1 – specificity.

**Supplementary Figure 10.**
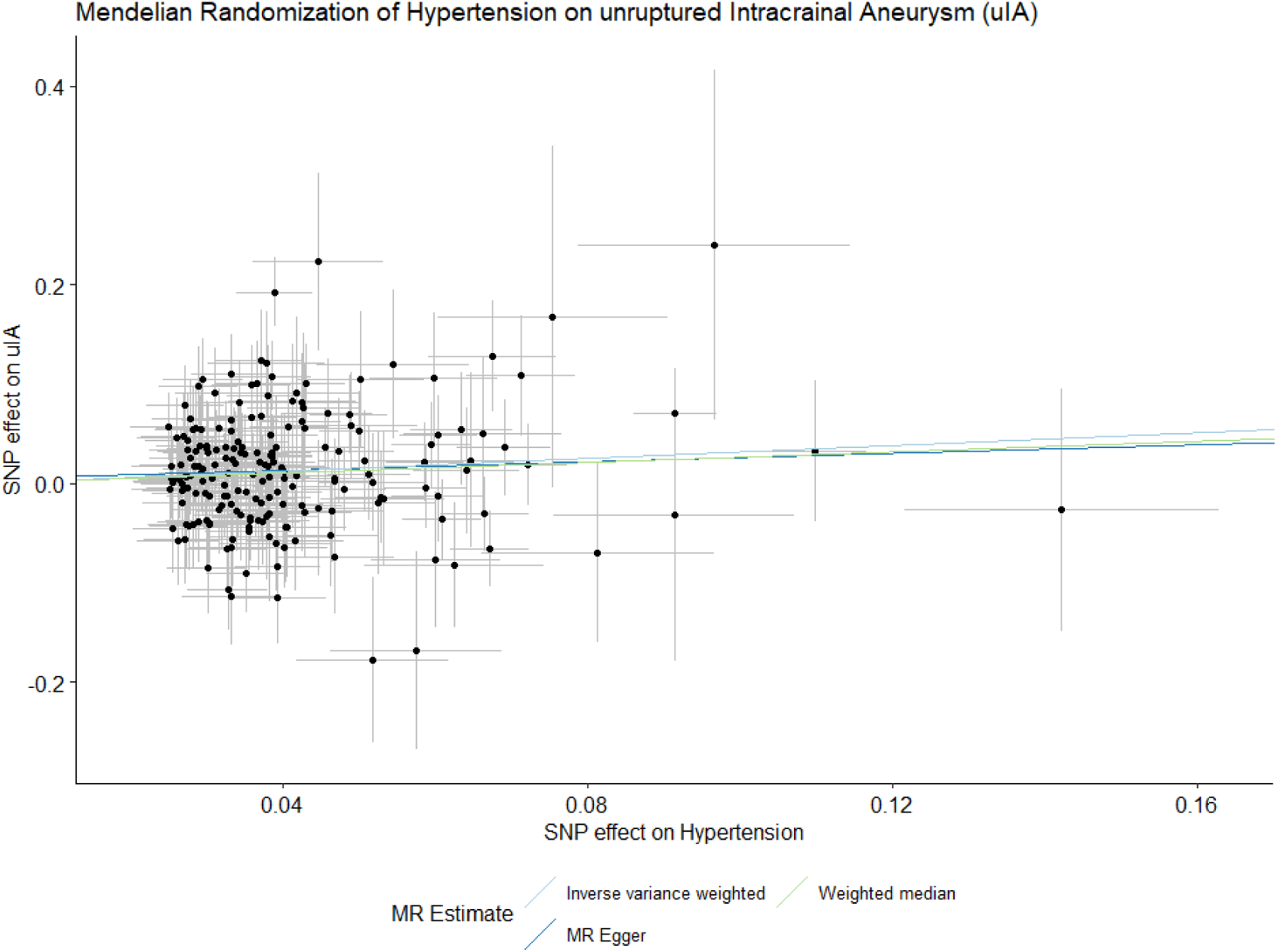
Mendelian randomization of hypertension on unruptured IA. Scatter-plot showing the effect estimate of each instrumental variable on hypertension (exposure) and risk of unruptured intracranial aneurysms (outcome) in the multiple instrumental variable Mendelian Randomization.

**Supplementary Figure 11.**
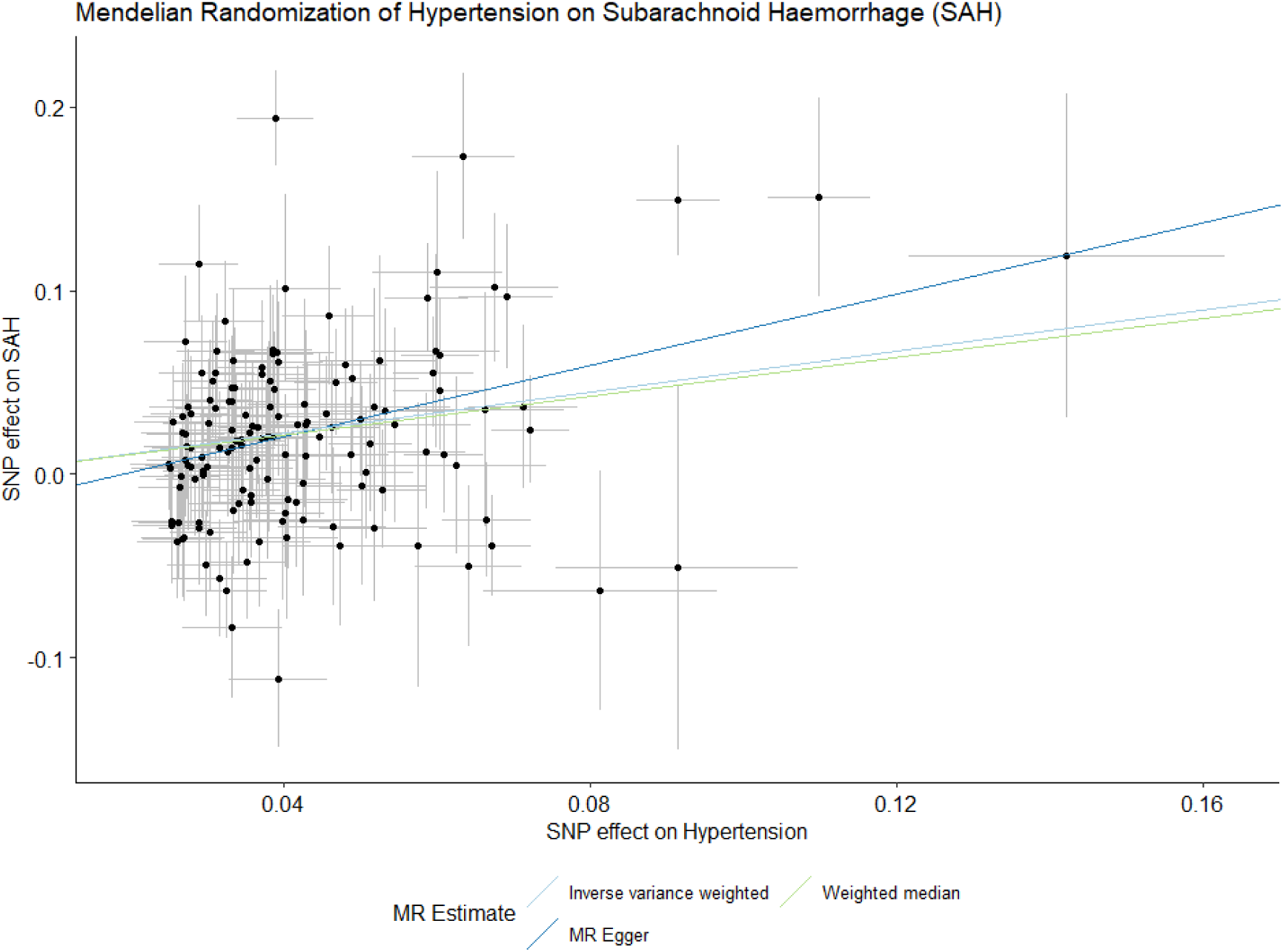
Mendelian randomization of hypertension on SAH. Scatter-plot showing the effect estimate of each instrumental variable on hypertension (exposure) and risk of subarachnoid haemorrhage (outcome) in the multiple instrumental variable Mendelian Randomization.

## Supplementary Tables

**Supplementary Table 1.**
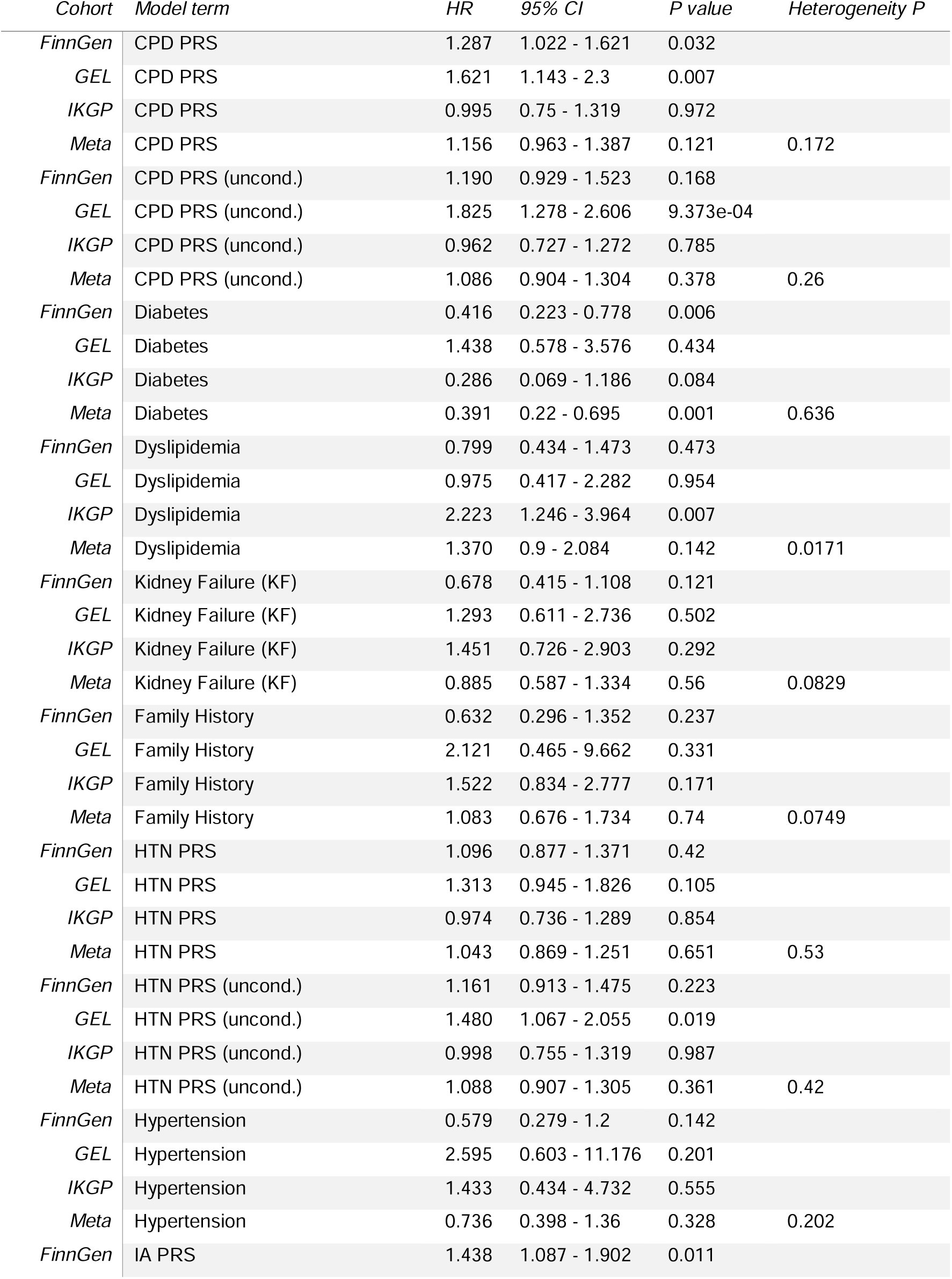
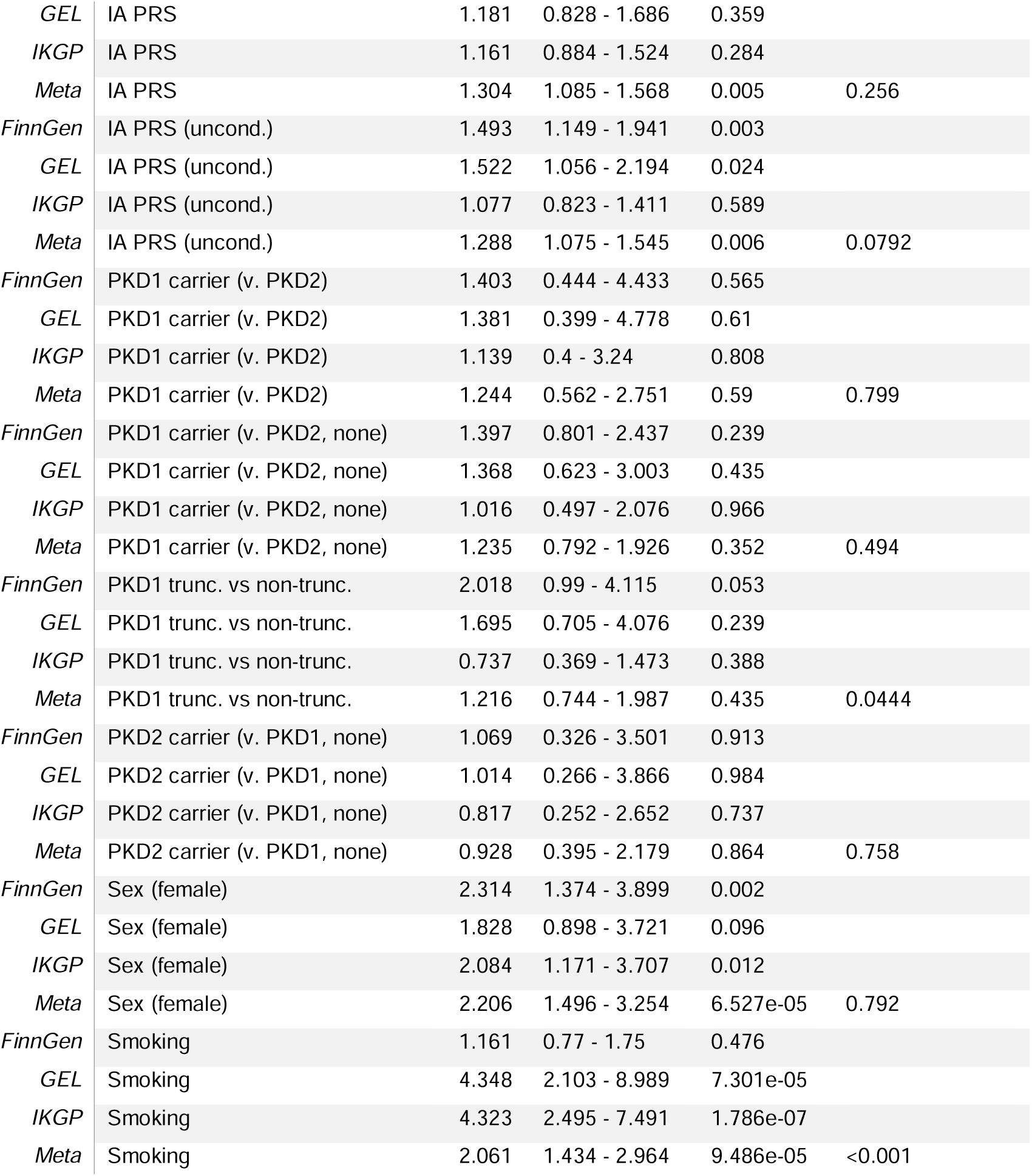
Results of univariate Cox regression models. Statistical results of univariate Cox regression models, with first 10 PCs as covariates. **Supplementary Table 1** | Results of univariate Cox regression models. Hazard ratios (HR) with 95% confidence intervals (CI) and *P* values for the association of each term with the combined endpoint of intracranial aneurysm or subarachnoid haemorrhage, estimated separately in the FinnGen, Genomics England (GEL) and Irish Kidney Gene Project (IKGP) cohorts. Each model was fitted univariately with the first 10 genetic principal components as covariates. Meta-analysed estimates were obtained by inverse-variance weighting across cohorts, with the accompanying heterogeneity *P* value testing for differences in effect between cohorts. CPD, cigarettes per day; HTN, hypertension; IA, intracranial aneurysm; PRS, polygenic risk score; uncond., polygenic score derived from unconditioned GWAS summary statistics.

**Supplementary Table 2.**
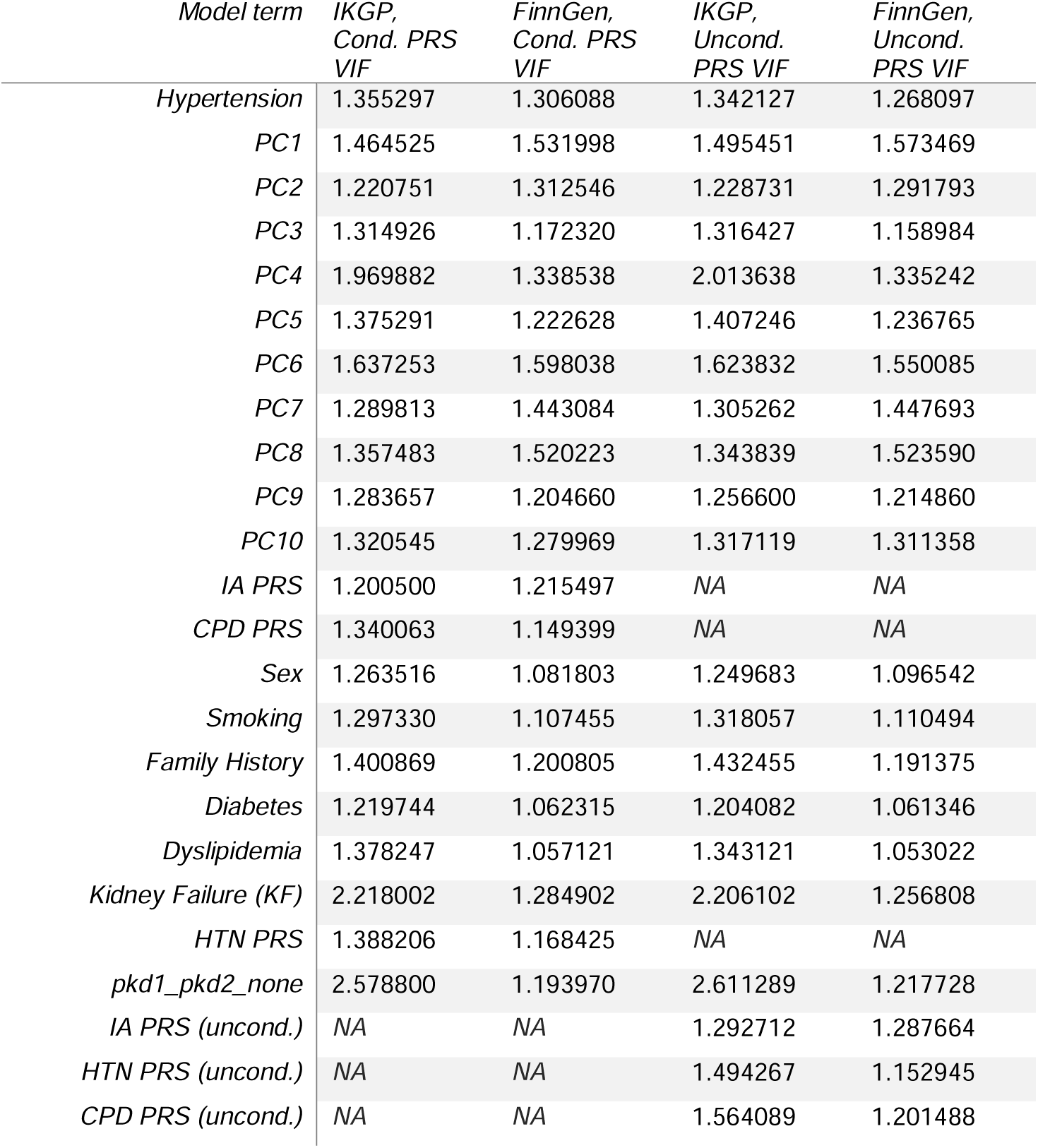
Variance inflation factors for multivariate Cox regression models. Variance inflation factors (VIF) for terms in multivariate Cox regression models, with first 10 PCs as covariates. **Supplementary Table 2** | Variance inflation factors for multivariate Cox regression models. Variance inflation factors (VIF) for each term in the multivariate Cox regression models fitted in the IKGP and FinnGen cohorts, using polygenic scores derived from either mtCOJO-conditioned or unconditioned GWAS summary statistics, with the first 10 genetic principal components as covariates. NA indicates that the term was not included in that model. CPD, cigarettes per day; HTN, hypertension; IA, intracranial aneurysm; PC, principal component; PRS, polygenic risk score.

**Supplementary Table 3.** Results of multivariate Cox regression models. Statistical results of multivariate Cox regression models, with first 10 PCs as covariates. **Supplementary Table 3** | Results of multivariate Cox regression models. Hazard ratios (HR) with 95% confidence intervals (CI) and *P* values for each term in the multivariate Cox regression models, fitted separately using polygenic scores derived from mtCOJO-conditioned (Cond. PRS) or unconditioned (Uncond. PRS) GWAS summary statistics, in the FinnGen, Genomics England (GEL) and Irish Kidney Gene Project (IKGP) cohorts. All models included the first 10 genetic principal components as covariates. Meta-analysed estimates were obtained by inverse-variance weighting across cohorts, with the accompanying heterogeneity *P* value testing for differences in effect between cohorts. CPD, cigarettes per day; HTN, hypertension; IA, intracranial aneurysm; PRS, polygenic risk score.

| *Cohort* | *Model* | *Model term* | *HR* | *95% CI* | *P* | *Heterogeneity P* |
| --- | --- | --- | --- | --- | --- | --- |
| *FinnGen* | Cond. PRS | CPD PRS | 1.324 | 0.99 - 1.77 | 0.058 |  |
| *GEL* | Cond. PRS | CPD PRS | 1.678 | 1.172 - 2.404 | 0.005 |  |
| *IKGP* | Cond. PRS | CPD PRS | 0.950 | 0.682 - 1.324 | 0.762 |  |
| *Meta* | Cond. PRS | CPD PRS | 1.155 | 0.933 - 1.429 | 0.186 | 0.133 |
| *FinnGen* | Uncond. PRS | CPD PRS (uncond.) | 1.188 | 0.864 - 1.633 | 0.289 |  |
| *GEL* | Uncond. PRS | CPD PRS (uncond.) | 1.790 | 1.245 - 2.574 | 0.002 |  |
| *IKGP* | Uncond. PRS | CPD PRS (uncond.) | 0.926 | 0.652 - 1.314 | 0.666 |  |
| *Meta* | Uncond. PRS | CPD PRS (uncond.) | 1.076 | 0.864 - 1.342 | 0.513 | 0.278 |
| *FinnGen* | Cond. PRS | Diabetes | 0.315 | 0.148 - 0.669 | 0.003 |  |
| *GEL* | Cond. PRS | Diabetes | 1.154 | 0.444 - 3 | 0.769 |  |
| *IKGP* | Cond. PRS | Diabetes | 0.502 | 0.105 - 2.404 | 0.388 |  |
| *Meta* | Cond. PRS | Diabetes | 0.345 | 0.173 - 0.688 | 0.003 | 0.601 |
| *FinnGen* | Uncond. PRS | Diabetes | 0.313 | 0.148 - 0.664 | 0.002 |  |
| *GEL* | Uncond. PRS | Diabetes | 1.267 | 0.483 - 3.323 | 0.631 |  |
| *IKGP* | Uncond. PRS | Diabetes | 0.509 | 0.107 - 2.415 | 0.395 |  |
| *Meta* | Uncond. PRS | Diabetes | 0.345 | 0.173 - 0.688 | 0.003 | 0.584 |
| *FinnGen* | Cond. PRS | Dyslipidemia | 0.609 | 0.29 - 1.28 | 0.191 |  |
| *GEL* | Cond. PRS | Dyslipidemia | 0.909 | 0.377 - 2.19 | 0.832 |  |
| *IKGP* | Cond. PRS | Dyslipidemia | 1.263 | 0.651 - 2.449 | 0.489 |  |
| *Meta* | Cond. PRS | Dyslipidemia | 0.910 | 0.557 - 1.487 | 0.707 | 0.148 |
| *FinnGen* | Uncond. PRS | Dyslipidemia | 0.624 | 0.294 - 1.324 | 0.219 |  |
| *GEL* | Uncond. PRS | Dyslipidemia | 0.931 | 0.383 - 2.262 | 0.875 |  |
| *IKGP* | Uncond. PRS | Dyslipidemia | 1.234 | 0.642 - 2.372 | 0.528 |  |
| *Meta* | Uncond. PRS | Dyslipidemia | 0.912 | 0.56 - 1.484 | 0.709 | 0.173 |
| *FinnGen* | Cond. PRS | Kidney Failure (KF) | 0.697 | 0.372 - 1.304 | 0.258 |  |
| *GEL* | Cond. PRS | Kidney Failure (KF) | 1.314 | 0.591 - 2.922 | 0.503 |  |
| *IKGP* | Cond. PRS | Kidney Failure (KF) | 1.246 | 0.571 - 2.722 | 0.581 |  |
| *Meta* | Cond. PRS | Kidney Failure (KF) | 0.867 | 0.537 - 1.402 | 0.561 | 0.25 |
| *FinnGen* | Uncond. PRS | Kidney Failure (KF) | 0.700 | 0.374 - 1.31 | 0.265 |  |
| *GEL* | Uncond. PRS | Kidney Failure (KF) | 1.254 | 0.562 - 2.794 | 0.581 |  |
| *IKGP* | Uncond. PRS | Kidney Failure (KF) | 1.258 | 0.575 - 2.751 | 0.566 |  |
| *Meta* | Uncond. PRS | Kidney Failure (KF) | 0.870 | 0.54 - 1.401 | 0.567 | 0.245 |
| *FinnGen* | Cond. PRS | Family History | 0.549 | 0.212 - 1.417 | 0.215 |  |
| *GEL* | Cond. PRS | Family History | 1.592 | 0.274 - 9.236 | 0.604 |  |
| *IKGP* | Cond. PRS | Family History | 1.352 | 0.711 - 2.573 | 0.358 |  |
| *Meta* | Cond. PRS | Family History | 0.982 | 0.586 - 1.646 | 0.945 | 0.101 |
| *FinnGen* | Uncond. PRS | Family History | 0.514 | 0.199 - 1.327 | 0.169 |  |
| *GEL* | Uncond. PRS | Family History | 1.439 | 0.235 - 8.801 | 0.694 |  |
| *IKGP* | Uncond. PRS | Family History | 1.358 | 0.708 - 2.605 | 0.356 |  |
| *Meta* | Uncond. PRS | Family History | 0.955 | 0.568 - 1.606 | 0.863 | 0.0781 |
| *FinnGen* | Cond. PRS | HTN PRS | 1.239 | 0.948 - 1.62 | 0.116 |  |
| *GEL* | Cond. PRS | HTN PRS | 1.307 | 0.92 - 1.859 | 0.135 |  |
| *IKGP* | Cond. PRS | HTN PRS | 0.994 | 0.709 - 1.395 | 0.974 |  |
| *Meta* | Cond. PRS | HTN PRS | 1.138 | 0.922 - 1.405 | 0.228 | 0.318 |
| *FinnGen* | Uncond. PRS | HTN PRS (uncond.) | 1.344 | 1.005 - 1.797 | 0.046 |  |
| *GEL* | Uncond. PRS | HTN PRS (uncond.) | 1.450 | 1.008 - 2.086 | 0.045 |  |
| *IKGP* | Uncond. PRS | HTN PRS (uncond.) | 0.992 | 0.704 - 1.399 | 0.966 |  |
| *Meta* | Uncond. PRS | HTN PRS (uncond.) | 1.196 | 0.966 - 1.48 | 0.1 | 0.174 |
| *FinnGen* | Cond. PRS | Hypertension | 0.544 | 0.239 - 1.24 | 0.148 |  |
| *GEL* | Cond. PRS | Hypertension | 2.445 | 0.554 - 10.782 | 0.238 |  |
| *IKGP* | Cond. PRS | Hypertension | 0.377 | 0.094 - 1.51 | 0.168 |  |
| *Meta* | Cond. PRS | 0 | 0.495 | 0.244 - 1.004 | 0.051 | 0.655 |
| *FinnGen* | Uncond. PRS | Hypertension | 0.520 | 0.231 - 1.172 | 0.115 |  |
| *GEL* | Uncond. PRS | Hypertension | 2.160 | 0.486 - 9.606 | 0.312 |  |
| *IKGP* | Uncond. PRS | Hypertension | 0.384 | 0.096 - 1.53 | 0.175 |  |
| *Meta* | Uncond. PRS | Hypertension | 0.481 | 0.239 - 0.968 | 0.04 | 0.711 |
| *FinnGen* | Cond. PRS | IA PRS | 1.325 | 0.959 - 1.832 | 0.088 |  |
| *GEL* | Cond. PRS | IA PRS | 1.052 | 0.728 - 1.519 | 0.788 |  |
| *IKGP* | Cond. PRS | IA PRS | 1.201 | 0.89 - 1.619 | 0.23 |  |
| *Meta* | Cond. PRS | IA PRS | 1.265 | 1.029 - 1.554 | 0.025 | 0.639 |
| *FinnGen* | Uncond. PRS | IA PRS (uncond.) | 1.420 | 1.023 - 1.972 | 0.036 |  |
| *GEL* | Uncond. PRS | IA PRS (uncond.) | 1.270 | 0.861 - 1.872 | 0.228 |  |
| *IKGP* | Uncond. PRS | IA PRS (uncond.) | 1.131 | 0.824 - 1.554 | 0.445 |  |
| *Meta* | Uncond. PRS | IA PRS (uncond.) | 1.285 | 1.041 - 1.586 | 0.019 | 0.293 |
| *FinnGen* | Cond. PRS | PKD1 carrier (v. PKD2, none) | 1.495 | 0.839 - 2.662 | 0.172 |  |
| *GEL* | Cond. PRS | PKD1 carrier (v. PKD2, none) | 1.499 | 0.627 - 3.581 | 0.362 |  |
| *IKGP* | Cond. PRS | PKD1 carrier (v. PKD2, none) | 0.699 | 0.247 - 1.982 | 0.501 |  |
| *Meta* | Cond. PRS | PKD1 carrier (v. PKD2, none) | 1.245 | 0.747 - 2.076 | 0.401 | 0.213 |
| *FinnGen* | Uncond. PRS | PKD1 carrier (v. PKD2, none) | 1.541 | 0.86 - 2.763 | 0.146 |  |
| *GEL* | Uncond. PRS | PKD1 carrier (v. PKD2, none) | 1.467 | 0.61 - 3.527 | 0.392 |  |
| *IKGP* | Uncond. PRS | PKD1 carrier (v. PKD2, none) | 0.680 | 0.238 - 1.937 | 0.47 |  |
| *Meta* | Uncond. PRS | PKD1 carrier (v. PKD2, none) | 1.265 | 0.756 - 2.116 | 0.372 | 0.182 |
| *FinnGen* | Cond. PRS | Sex (female) | 2.240 | 1.232 - 4.073 | 0.008 |  |
| *GEL* | Cond. PRS | Sex (female) | 2.017 | 0.958 - 4.245 | 0.065 |  |
| *IKGP* | Cond. PRS | Sex (female) | 1.716 | 0.896 - 3.283 | 0.103 |  |
| *Meta* | Cond. PRS | Sex (female) | 1.985 | 1.283 - 3.072 | 0.002 | 0.552 |
| *FinnGen* | Uncond. PRS | Sex (female) | 2.374 | 1.306 - 4.314 | 0.005 |  |
| *GEL* | Uncond. PRS | Sex (female) | 1.879 | 0.887 - 3.98 | 0.1 |  |
| *IKGP* | Uncond. PRS | Sex (female) | 1.646 | 0.864 - 3.138 | 0.13 |  |
| *Meta* | Uncond. PRS | Sex (female) | 2.007 | 1.296 - 3.107 | 0.002 | 0.414 |
| *FinnGen* | Cond. PRS | Smoking | 1.569 | 0.924 - 2.664 | 0.096 |  |
| *GEL* | Cond. PRS | Smoking | 4.628 | 2.166 - 9.89 | 7.665e-05 |  |
| *IKGP* | Cond. PRS | Smoking | 6.272 | 3.335 - 11.795 | 1.216e-08 |  |
| *Meta* | Cond. PRS | Smoking | 2.847 | 1.881 - 4.307 | 7.400e-07 | 0.00117 |
| *FinnGen* | Uncond. PRS | Smoking | 1.566 | 0.925 - 2.652 | 0.095 |  |
| *GEL* | Uncond. PRS | Smoking | 4.688 | 2.172 - 10.12 | 8.288e-05 |  |
| *IKGP* | Uncond. PRS | Smoking | 6.435 | 3.406 - 12.159 | 9.794e-09 |  |
| *Meta* | Uncond. PRS | Smoking | 2.864 | 1.889 - 4.34 | 7.093e-07 | <0.001 |

**Supplementary Table 4.** Comparison of models for all cohorts. Comparison between nested models for all cohorts with the likelihood ratio test. **Supplementary Table 4** | Comparison of nested models within each cohort. Likelihood ratio tests comparing nested Cox regression models (Base clinical, Base + monogenic, Base + PRS, Base + monogenic + PRS) in the FinnGen, Genomics England (GEL) and Irish Kidney Gene Project (IKGP) cohorts. *n* denotes the number of participants included in each comparison. PRS, polygenic risk score.

| *Model* | *Cohort* | *n* | *P* |
| --- | --- | --- | --- |
| *Base Clinical Vs Base + Monogenic* | FinnGen | 574 | 0.290 |
| *Base Clinical Vs Base + Prs* | FinnGen | 574 | 0.004 |
| *Base + Monogenic Vs Base + Monogenic + Prs* | FinnGen | 574 | 0.004 |
| *Base + Prs Vs Base + Monogenic + Prs* | FinnGen | 574 | 0.336 |
| *Base Clinical Vs Base + Monogenic* | IKGP | 461 | 0.726 |
| *Base Clinical Vs Base + Prs* | IKGP | 461 | 0.633 |
| *Base + Prs Vs Base + Monogenic + Prs* | IKGP | 461 | 0.802 |
| *Base + Monogenic Vs Base + Monogenic + Prs* | IKGP | 461 | 0.678 |
| *Base Clinical Vs Base + Monogenic* | GEL | 895 | 0.614 |
| *Base + Monogenic Vs Base + Monogenic + Prs* | GEL | 895 | 0.008 |
| *Base Clinical Vs Base + Prs* | GEL | 895 | 0.009 |
| *Base + Prs Vs Base + Monogenic + Prs* | GEL | 895 | 0.534 |

**Supplementary Table 5.** Meta analysis of c-index across cohorts. Inverse-variance weighted meta-analysis of c-index across three cohorts for each model. **Supplementary Table 5** | Meta-analysis of the c-index across cohorts. Inverse-variance weighted meta-analysis of Harrell’s concordance index (c-index) with 95% confidence intervals (CI) for each model across the FinnGen, Genomics England and Irish Kidney Gene Project cohorts. The heterogeneity *P* value tests for differences in discrimination between cohorts. PRS, polygenic risk score.

| *Model* | *C-index* | *95% CI* | *Heterogeneity*<br>*P* |
| --- | --- | --- | --- |
| *Base + PRS* | 0.821 | 0.708 - 0.897 | 4.570e-05 |
| *Base + monogenic* | 0.808 | 0.671 - 0.896 | 4.167e-06 |
| *Base + monogenic + PRS* | 0.825 | 0.717 - 0.898 | 7.816e-05 |
| *Base clinical* | 0.804 | 0.662 - 0.896 | 1.609e-06 |

